# Multivariate genome-wide association meta-analysis of over 1 million subjects identifies loci underlying multiple substance use disorders

**DOI:** 10.1101/2022.01.06.22268753

**Authors:** Alexander S. Hatoum, Sarah M.C. Colbert, Emma C. Johnson, Spencer B. Huggett, Joseph D. Deak, Gita Pathak, Mariela V. Jennings, Sarah E. Paul, Nicole R. Karcher, Isabella Hansen, David A.A. Baranger, Alexis Edwards, Andrew Grotzinger, Substance Use Disorder Working Group of the Psychiatric Genomics Consortium, Elliot M. Tucker-Drob, Henry R. Kranzler, Lea K. Davis, Sandra Sanchez-Roige, Renato Polimanti, Joel Gelernter, Howard J. Edenberg, Ryan Bogdan, Arpana Agrawal

**Author notes:** Please send all correspondence to: Washington University School of Medicine, Department of Psychiatry, 660 S. Euclid, CB 8134, Saint Louis, MO 63110, USA. These authors jointly supervised the research and contributed equally. The full list of the members of the Psychiatric Genomics Consortium’s Substance Use Disorders (PGC-SUD) Working Group is in the appendix. [how we list authors depends on journal]. **Author contributions** ASH designed the study and conducted analyses. SMCC, ECJ, SBH, JDD, GP, MVJ, SS-R, SEP, NRK, IH and DAAB conducted various analyses. AA, RB, HJE, and JG supervised the study. AG and ETD provided statistical guidance. AE, HRK, RP, LKD, SS-R guided interpretation of key findings. ASH, HJE, JG, RB, and AA drafted the manuscript. ASH, SMCC, JDD, MVJ, SEP, NRK, and IH, organized the data. The PGC-SUD consortium members provided insight into various aspects of analyses and interpretation. All named authors reviewed, edited and approved the submission. **Disclosure:** Dr. Kranzler is a member of advisory boards for Dicerna Pharmaceuticals and Sophrosyne Pharmaceuticals, and Enthion Pharmaceuticals; a consultant to Sobrera Pharmaceuticals; and a member of the American Society of Clinical Psychopharmacology’s Alcohol Clinical Trials Initiative, which was supported in the last three years by Alkermes, Dicerna, Ethypharm, Lundbeck, Mitsubishi, and Otsuka. Drs. Kranzler and Gelernter are named as inventors on PCT patent application #15/878,640 entitled: “Genotype-guided dosing of opioid agonists,” filed January 24, 2018. Publicly available data were also taken from the psychiatric genomics consortium: https://www.med.unc.edu/pgc/, and the GSCAN consortium: https://conservancy.umn.edu/handle/11299/201564. **Funding:** T32DA007261 (ASH), DA54869 (AA, JG, HE), DA54750 (AA, RB), K02DA32573 (AA), R21AA027827 (RB), K01DA51759 (ECJ), DP1DA54394 (SS-R), R01AA027522 (AE), F31AA029934 (SEP), R01MH120219 (EMTD, ADG), RF1AG073593 (EMTD, ADG), P30AG066614 (EMTD), P2CHD042849 (EMTD), R33DA047527 (RP, GAP), T32 AA028259 (JDD).

## Abstract

Genetic liability to substance use disorders can be parsed into loci conferring general and substance-specific addiction risk. We report a multivariate genome-wide association study that disaggregates general and substance-specific loci for problematic alcohol use, problematic tobacco use, and cannabis and opioid use disorders in a sample of **1,025,550** individuals of European and **92,630 individuals of** African descent. Nineteen loci were genome-wide significant for the general addiction risk factor (*addiction-rf*), which showed high polygenicity. Across ancestries *PDE4B* was significant (among others), suggesting dopamine regulation as a cross-trait vulnerability. The *addiction-rf* polygenic risk score was associated with substance use disorders, psychopathologies, somatic conditions, and environments associated with the onset of addictions. Substance-specific loci (9 for alcohol, 32 for tobacco, 5 for cannabis, 1 for opioids) included metabolic and receptor genes. These findings provide insight into the genetic architecture of general and substance-specific use disorder risk that may be leveraged as treatment targets.

---

The lives lost, impacts on individuals and families, and socioeconomic costs attributable to substance use reflect a growing public health crisis^1^. For example, in the United States, 13.5% of deaths among young adults^2^ are attributable to alcohol, smoking is the leading risk factor for mortality in males^3^, and the odds of dying by opioid overdose are greater than those of dying in a motor vehicle crash^4^. Despite the large impact of substance use and disorders^5^, there is limited knowledge of the molecular genetic underpinnings of addiction broadly. This may be partially attributable to the often-siloed study of substance use disorders (SUDs) as unique substance-specific entities, despite shared symptomatology and widespread comorbidity among SUDs.

Individual SUDs are heritable (h^2^ ∼ 50-60%) and highly polygenic^6,7^. Recent large-scale genome-wide association studies (GWAS) have identified loci associated with problematic drinking^8,9^, alcohol use disorder (AUD)^10,11^, cigarettes smoked per day^12^, nicotine dependence^13,14^, cannabis use disorder (CUD)^15^ and opioid use disorder (OUD)^16^. Echoing evidence from twin and family studies^17^, these GWAS show that the genomic architecture of SUDs is characterized by a high degree of commonality^18^. This shared genetic architecture is correlated with genetic liability to non-substance psychopathology (e.g., major depression, schizophrenia) and traits such as executive functioning, risk-taking and neuroticism that underlie neurobiological models of addiction^18^. Even after accounting for genetic correlations with non-problematic substance use and with other psychiatrically-relevant traits and disorders, there is considerable variance that is unique to a general risk for addiction, indicating that a liability to addiction reflects more than just the combined genetic liability to substance use and psychopathology^18-20^.

Characterizing the genetic architecture associated with a liability to SUDs in general and to specific SUDs could expedite advances in the nosology, prevention, and treatment of these disorders. To this end, we conducted a multivariate GWAS of largest available discovery GWAS of substance use disorders, including problematic alcohol use (PAU: N=435,563)^8^, problematic tobacco use (PTU: N=270,120)^12,14,18^, cannabis use disorder (CUD: N=384,032)^15^ and opioid use disorder (OUD: N=79,729)^16^. *First*, we partitioned SNP effects into 5 sources of variation: ***(1)*** a general addiction risk factor (referred to as *addiction-rf*), and risks specific to ***(2)*** alcohol, ***(3)*** nicotine, ***(4)*** cannabis and ***(5)*** opioids. *Second*, we identified biological pathways underlying risk for these 5 SUD phenotypes using gene, eQTL, and pathway enrichment analyses. *Third,* we examined whether currently available medications could potentially be repurposed to treat SUDs by examining whether the gene expression profiles of *in vitro* neuronal cell lines treated with these chemical compounds^21^ are associated with gene expression profiles in postmortem brain tissue correlated with *addiction-rf. Fourth*, we assessed the association of a polygenic risk score (PRS) derived from *addiction-rf* with general and specific SUD phenotypes in an independent case/control sample. *Fifth,* we examined the extent to which genetic liability to *addiction-rf* is shared with other phenotypes (e.g., physical and mental health outcomes) by estimating genetic correlations with published GWAS of those traits. We also examined genetic evidence for causality for phenotypes that were significantly genetically correlated with *addiction-rf. Sixth*, we tested whether the *addiction-rf* PRS was associated with: (a) medical diagnoses derived from electronic medical records in a population cohort (BioVU, n=66,914)^22^, and (b) 1,480 behavioral and neural phenotypes in largely substance-naïve 9-10-year-old children (n=4,490) from the Adolescent Brain and Cognitive Development (ABCD) Study^®23^.

## Results

### European ancestry GWAS: Addiction risk factor

As shown in our prior study^18^, the single factor model fit the data well (*X*^2^(1) = .017, p = 0.895, CFI = 1, SRMR = 0.002; factor loadings 0.36 – 0.93; lowest for PTU; **Supplemental Methods** for full model). Using genomic structural equation modeling^24^, we identified 19 independent (r2 < 0.1) genome-wide significant (GWS) SNPs that map onto 17 genomic risk loci that were associated with the *addiction-rf* (**Figure 1; Table 1; Supplemental Table 1** for independent SNPs and **Supplemental Table 2** for genomic risk loci). The most significant SNP (rs6589386, p=2.9e-12) was intergenic, but closest to *DRD2* (Dopamine receptor 2), which was GWS in gene-based analyses (p=7.9e-12). Further, rs6589386 was an expression quantitative trait locus (eQTL) for *DRD2* in the cerebellum, and Hi-C analyses (in FUMA)^25^ revealed that the variant made chromatin contact with the promoter of the gene (**Supplemental Figure 1**).

**Figure 1.**
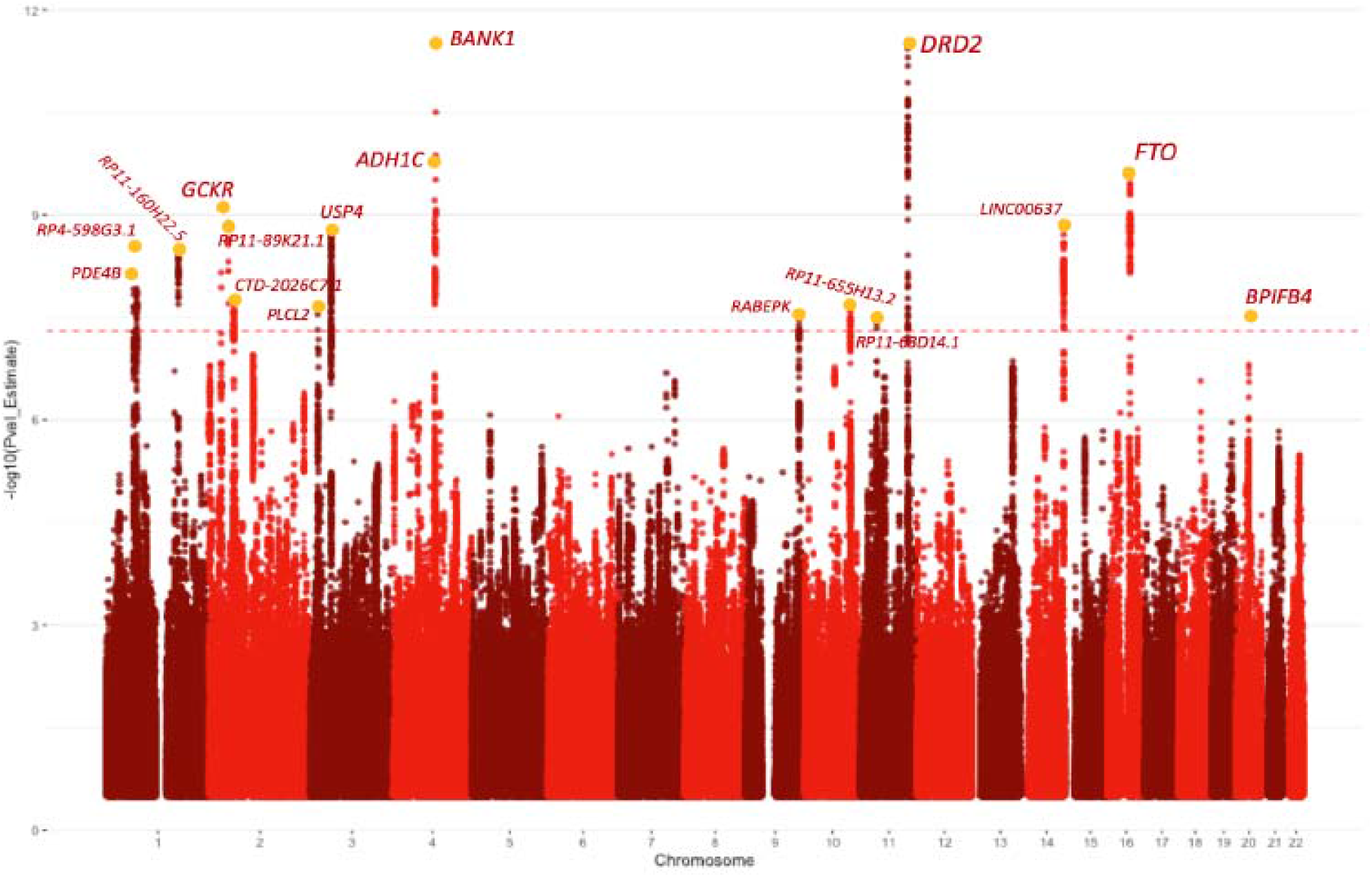
Manhattan plot of signals associated with the *addiction-rf*, a general genetic factor underlying problematic alcohol use, problem tobacco use, cannabis use disorder and opioid use disorder. Each SNP peak is annotated with the closest gene to a fine-mapped SNP with a posterior probability of inclusion of 1 or (if none) the closest gene to the lead SNP.

**Table 1.** Genome-wide significant risk loci for *addiction-rf*. Candidate SNPs = number of SNPs tagged. GWAS SNPs = variants related to that independent SNP. Chr = chromosome, A1 = effect allele.

| rsID | chr | A1 | position | European Ancestry |  |  |  | African Ancestry |  | Cross-ancestry |  |
| --- | --- | --- | --- | --- | --- | --- | --- | --- | --- | --- | --- |
|  |  |  |  | GSEM Beta<br>(effect allele) | p | Candidate<br>SNPS | GWAS<br>SNPs | Beta<br>(effect<br>allele) | p | Beta<br>(effect<br>allele) | p |
| rs1937455 | 1 | A | 66416939 | 0.013 | 7.74E-09 | 97 | 43 | 0.024 | 3.00E-03 | 0.013 | 0.100 |
| rs1475064 | 1 | A | 73882478 | 0.014 | 3.06E-09 | 312 | 161 | 0.09 | 6.25E-03 | 0.011 | 0.051 |
| rs2860846 | 1 | T | 174075924 | 0.014 | 3.37E-09 | 236 | 36 | NA | NA | NA | NA |
| rs1260326 | 2 | T | 27730940 | -0.015 | 7.60E-10 | 18 | 10 | NA | NA | NA | NA |
| rs570436 | 2 | C | 45142673 | -0.015 | 1.31E-09 | 50 | 8 | NA | NA | NA | NA |
| rs2717054 | 2 | G | 58046683 | -0.014 | 1.97E-08 | 229 | 44 | NA | NA | NA | NA |
| rs55855024 | 3 | C | 16850764 | -0.014 | 2.37E-08 | 71 | 35 | NA | NA | NA | NA |
| rs6795772 | 3 | C | 49365269 | 0.014 | 1.58E-09 | 463 | 265 | NA | NA | NA | NA |
| rs3114045 | 4 | T | 100252560 | -0.023 | 1.85E-10 | 278 | 134 | -0.019 | 0.554 | -0.012 | 2.60E-15 |
| rs1813006 | 4 | G | 103001649 | 0.037 | 3.18E-12 | 28 | 11 | NA | NA | NA | NA |
| rs864882 | 9 | C | 127968109 | 0.015 | 3.41E-08 | 107 | 26 | NA | NA | NA | NA |
| rs7073987 | 10 | C | 110565868 | 0.015 | 2.04E-08 | 126 | 61 | NA | NA | NA | NA |
| rs2861190 | 11 | C | 38517941 | -0.014 | 3.66E-08 | 186 | 117 | NA | NA | NA | NA |
| rs6589386 | 11 | C | 113443753 | 0.017 | 2.92E-12 | 132 | 65 | NA | NA | NA | NA |
| rs10083370 | 14 | G | 104314182 | 0.014 | 1.53E-09 | 249 | 89 | NA | NA | NA | NA |
| rs28567725 | 16 | T | 53826028 | 0.016 | 2.50E-10 | 116 | 83 | 0.012 | .457 | 0.007 | 6.49E-12 |
| rs2424952 | 20 | T | 31685873 | 0.030 | 3.21E-08 | 12 | 4 | NA | NA | NA | NA |

**Table 2.**
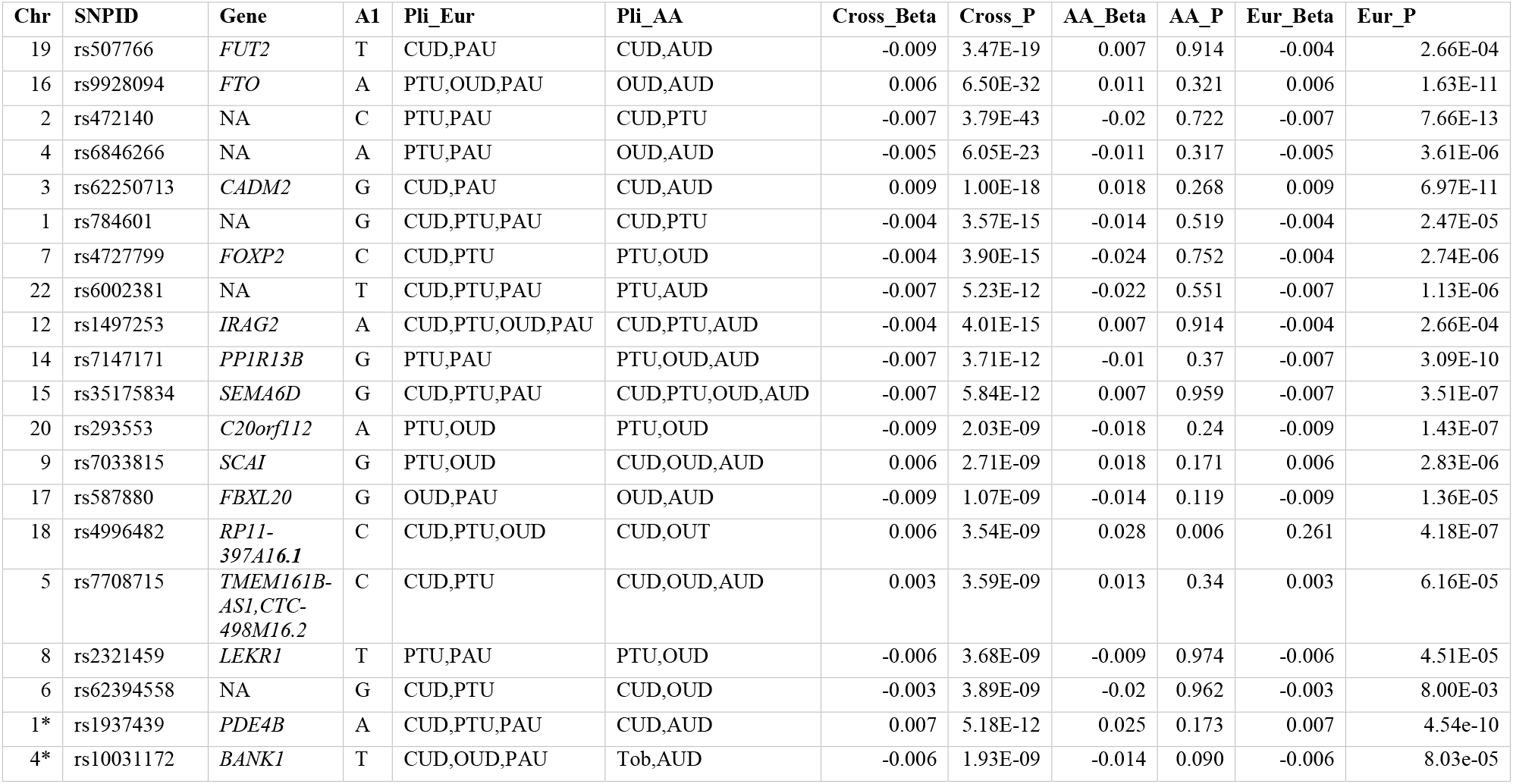
Results from Cross-Ancestry Meta-Analysis. Results from a random effects cross-ancestry meta-analysis conducted across the European and African ancestry samples with METAFOR. Significant top results from each chromosome are shown. European and African ancestry results were estimated with ASSET. Loci with any pleiotropic (pairwise or more) effect was included for each ancestral group. We took the top loci from each chromosome. We then searched for any loci that overlap with genes implicated in the European GWAS, noted by a * in the chromosome column. The individual betas from the input and the cross-ancestry beta are also shown and those with consistent effects across all ancestries and in the cross ancestry meta-analysis are highlighted in yellow. Chr=Chromosome, SNPID=rsid, GENE= the gene name if the SNP was on a gene (otherwise NA), Pli_Eur = Traits loci is pleiotropic for in Europeans, Pli_AA = Traits loci is pleiotropic for in Africans, Cross_Beta = Beta in cross ancestry analysis, Cross_P = P-value in Cross-Ancestry analysis, AA_Beta = Beta in Arican Ancestry, AA_P = P-value in Arican Ancestry, Eur_Beta = Beta in European ancestry analysis, Eur_P = Pvalue in European Ancestry analysis. CUD=Cannabis Use Disorder, PAU = Problematic Alcohol Use (European ancestry only), AUD = Alcohol Use Disorder (African ancestry only), PTU = Problematic Tobacco Use, OUD = Opioid Use Disorder.

Gene-based analyses identified 42 significant associations (**Supplemental Table 3**) with the most significant signals in *FTO* (p=1.86E-13*), DRD2* (p*=*7.9e-12), and *PDE4B* (p=9.63E-11). Fine-mapping identified 123 GWS SNPs (of 660 non-independent GWS SNPs) in credible sets as potential causal SNPs based on the posterior probability of inclusion (**Supplemental Table 4**). Mapping the lead independent SNPs in the credible sets to their nearest gene based on posterior probability of 1, the following SNPs showed the strongest causal potential: rs1937455 (*PDE4B*), rs3739095 (*GTF3C2*), rs6718128 (*ZNF512*), rs4143308 (*RP11-89K21.1*), rs4953152 (*SIX3*), rs41335055 (*CTD-2026C7.1*), rs2678900 (*VRK*), rs7620024 (*TCTA*), rs283412 (*ADH1C*), rs901406 (*BANK1*), rs359590 (*RABEPK*), rs10083370 (*LINC00637*), rs1477196 (*FTO*), rs291699 (*CDK5RAP1*) (**Supplemental Table 4** and **Figure 1**). Pathway analysis of gene-based results revealed several significant GO terms including double-stranded DNA binding (p_bonferroni_=0.005), sequence-specific double-stranded DNA binding (p_bonferroni_=0.01), regulation of nervous system development (2 terms: p_bonferroni_=0.011 – 0.037), and positive regulation of transcription by RNA polymerase (p_bonferroni_=0.038) (**Supplemental Table 5**).

### European ancestry GWAS: Substance-specific risk

Q-SNP analysis^24^ indicated that some SNPs were associated individually with PAU, PTU, CUD and OUD, and that these associations were inconsistent with the effect of these SNPs via the *addiction-rf*. Many of these were top loci from past GWAS of individual SUDs that could be mechanistically linked to specific substances (e.g., *ADH1B* and alcohol, *CHRNA5* and tobacco; **Supplemental Figure 2A**). To assess whether loci were associated with only one substance, we used ASSET (1-sided *p* < 5e-8)^26^. Only SNPs that were associated at GWS with one of the individual substances in ASSET were considered as substance-specific. (**Supplemental Figure 2B-E**).

#### Problematic Alcohol Use

There were 9 independent SNPs in 6 loci associated specifically with PAU in ASSET (**Supplemental Figure 2B; Supplemental Table 6** and **7**). The top signal was rs1229984 in *ADH1B* (p-value=4.11E-68), as expected^7^. Gene-based enrichment analyses also implicated the alcohol dehydrogenase activity zinc dependent pathway (p_bonferroni_=0.0347; **Supplemental Table 8**). Several SNPs were in genes that metabolize sugars including *KLB* (rs13129401, p=2.37e-18), *GCKR* (rs1260326, *p*=1.53e-18), but the pathway analysis did not discover any particular metabolic pathway.

#### Problematic Tobacco Use

PTU was specifically associated with 32 independent SNPs in 12 loci. The top SNP was rs10419203 (p=5.12e-267), though the signal is likely driven by the *CHRNA5* missense variant, rs16969968 (p=2.79e-175), which has previously been linked to tobacco use (**Supplemental Figure 2C; Supplemental Table 9** and **10**). Several other SNPs were closest to genes encoding nicotinic acetylcholine receptors, including *CHRNA4, CHRNB4, CHRNB3, CHRNB2.* Gene-based enrichment implicated multiple pathways and gene sets related to nicotinic acetylcholine receptors (**Supplemental Table 11**). Specific dopamine-related associations were also noted (e.g., *PDE1C:* rs415820; p=2.35e-18; *DBH:* rs1108581;p=1.00e-14).

#### Cannabis Use Disorder

ASSET identified 5 substance-specific loci for CUD (**Supplemental Table 12** and **13),** with lead signals at rs11913634 (p=1.20e-15), rs8104317 (p=1.17e-13), rs72818514 (p=1.57e-09), rs11778040 (p=1.77e-09), and rs11715758 (p=4.84e-08; **Supplemental Figure 2D**). Interestingly, the lead signal rs11913634 (p = 1.20e-15) on chromosome 5 mapped to *FAM19A5*, a brain specific gene influencing histone modification, rs8104317 (p=1.17e13) mapped to *CACNA1A*, while rs11778040 mapped to the previously^15^ discovered signal for CUD near *CHRNA2 and EPHX2*. CUD-specific signals showed no significant gene-based enrichment.

#### Opioid Use Disorder

The only significant substance-specific signal for OUD was the well-characterized mu opioid receptor (*OPRM1*) SNP, rs1799971 (p=1.63e-08; Figure 2E). Gene-based analyses produced no significant findings.

**Figure 2.**
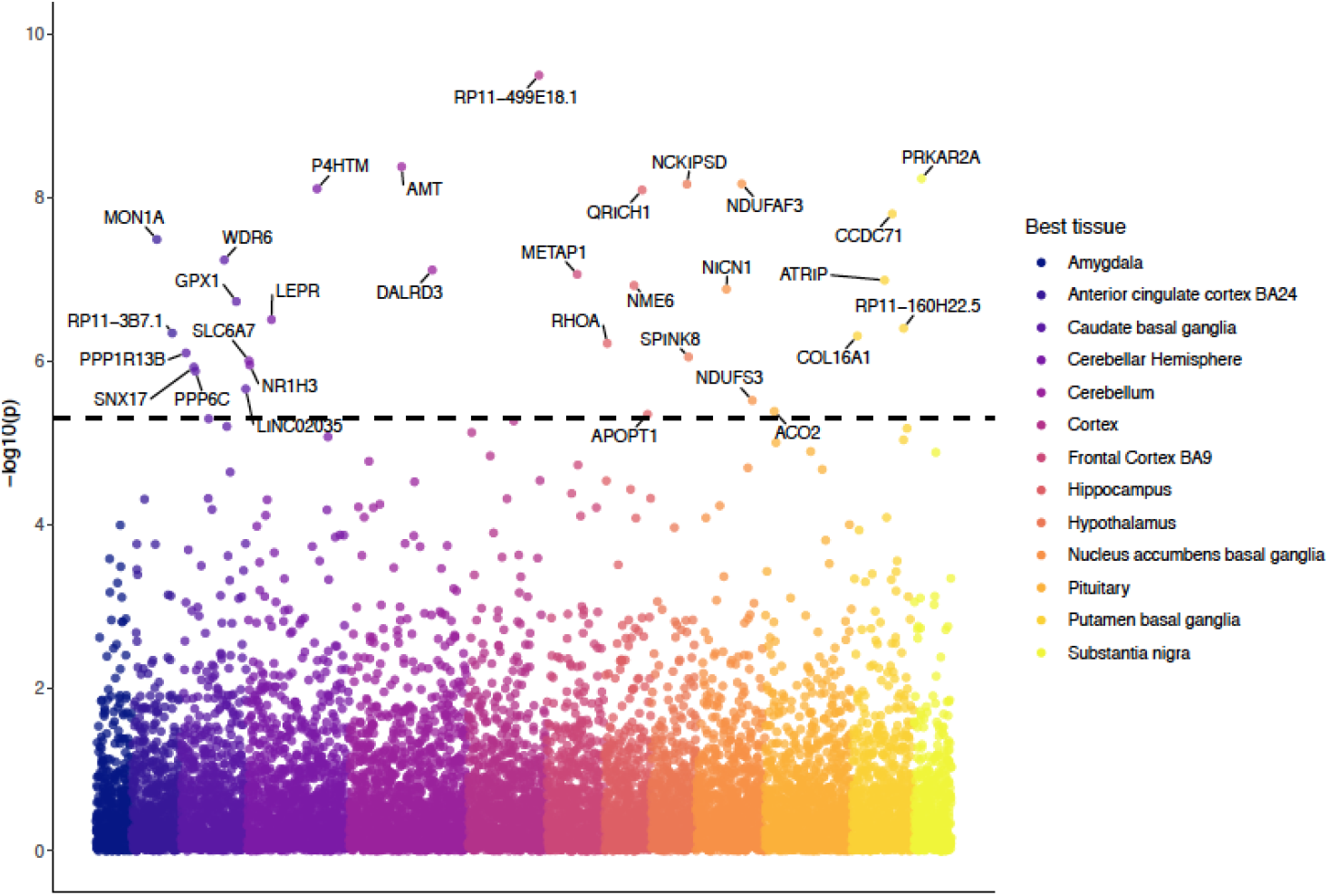
A Transcriptome-Wide Association Study (TWAS) of the *addiction-rf*, plotted as a Manhattan plot. Analysis were conducted in MetaXcan. The y-axis represents the log of the p-value, the color of the data point represents the tissue in which correlation between gene expression and outcome was the highest.

### African ancestry GWAS

The ASSET-based meta-analysis of GWAS data for AUD (N=82,705)^11^, tobacco dependence (based on the Fagerstrom Test for Nicotine Dependence, N=9,925)^14^, CUD (N=9,745)^15^, and OUD (N=32,088)^16^ in individuals of African ancestry yielded only 1 GWS SNP: rs2066702, an *ADH1B* variant that has been associated with alcohol-specific effects (**Supplemental Figure 3A**). One SNP, rs77193269 (p=4.92e-8), was GWS for AUD and tobacco dependence when considering ASSET loci pleiotropic for 2 traits (**Supplemental Figure 3B**). Due to its location, this SNP likely represents the *ADH1B* locus.

### Cross-Ancestry GWAS Results

We conducted a random-effects meta-analysis across ancestries using METASOFT^27^, with ancestry accounted for as a random effect. Only 317,447 SNPs were included after filtering for SNPs that did not show pleiotropic effects in ASSET (i.e., substance-specific) for either ancestry and for SNPs with genome-wide Cochran Q statistics indicating heterogeneity between the two ancestral groups. We found 68 SNPs (**Supplemental Figure 4**), which were challenging to map to lead loci due to the differences in LD structure across ancestries. **Table 2** lists SNP with lowest GWAS p-value on each chromosome. The most significant association was noted near the *FUT2* gene (rs507766, p = 3.47e-19). Many GWS signals were consistent with genes found in the European GWAS, including *FTO* (rs9928094, p = 6.50e-32) and *PDE4B* (rs1937439, p = 8.56e-12). We also identified two SNPs in genes which have previously been implicated in SUDs including *CADM2* (rs62250713, p=1.00E-18) and *FOXP2* (rs4727799, p=3.90E-15), both of which were within r^2^ = 0.6 of lead signals from the European GWAS.

### Polygenic architecture and power

The *addiction-rf* showed a highly polygenic architecture and smaller effect sizes per SNP (**Supplemental Figure 5A** and **5B**) than the original GWAS of PAU, PTU, CUD and OUD. In line with the discovery of variants of relatively large effects acting in PTU, OUD, and PAU (e.g., *CHRNA5, OPRM1,* and *ADH1B*)^6^, the substance-specific genetic architectures were relatively less polygenic.

### Transcriptome-wide Association Analysis and Drug Repurposing

A transcriptome-wide association analysis^28^ of *addiction-rf* identified 35 genes in 13 brain tissues (**Figure 2**; **Supplemental Table 14**). In a gene-set analysis using FUMA^25^, these genes were enriched for gene sets and pathways related to neural cells and T-cell processes (**Supplemental Figure 6**). Linking transcriptome-wide patterns to perturbagens that cross the blood-brain barrier from the Library of Integrated Network-Based Cellular Signatures (LINCS)^29^ database identified 104 medications approved by the U. S. Food and Drug Administration **(Supplemental Table 15)**. The top hits (p_bonferroni_ <0.0001) that reversed the transcriptional profile associated with the *addiction-rf* factor included mifepristone (rank #1), a progesterone blocker; varenicline (rank #18), a partial nicotinic acetylcholine receptor agonist drug used in smoking cessation; riluzole (rank #24), a glutamate release and voltage-dependent sodium channel inhibitor; and reboxetine (rank #14), a selective norepinephrine/noradrenaline reuptake inhibitor.

### LD Score Regression and Genetic Correlations

After Bonferroni correction (p < .05/1,547 =3.20e-5), the *addiction-rf* was genetically correlated with 250 phenotypes (**Figure 3; Supplemental Table 16**). Notably, 38 of these included somatic diseases that are often associated with one specific drug (e.g., lung cancer with tobacco, and pain-related conditions with opioids). As expected, we found significant genetic correlations between the *addiction-rf* and serious, trans-diagnostic psychopathologic behaviors, including suicide attempt (rg=.618, p=2.89e-33) and self-medication (e.g., using non-prescribed drugs or alcohol for anxiety, rg=.635, p=3.18e-5). The *addiction-rf* was correlated with, but remained separable based on 95% confidence intervals (rg=0.63,±.037, p=2.33e-231), from an externalizing factor^24^ that included similar indices of problematic substance use and behavioral measures.

**Figure 3.**
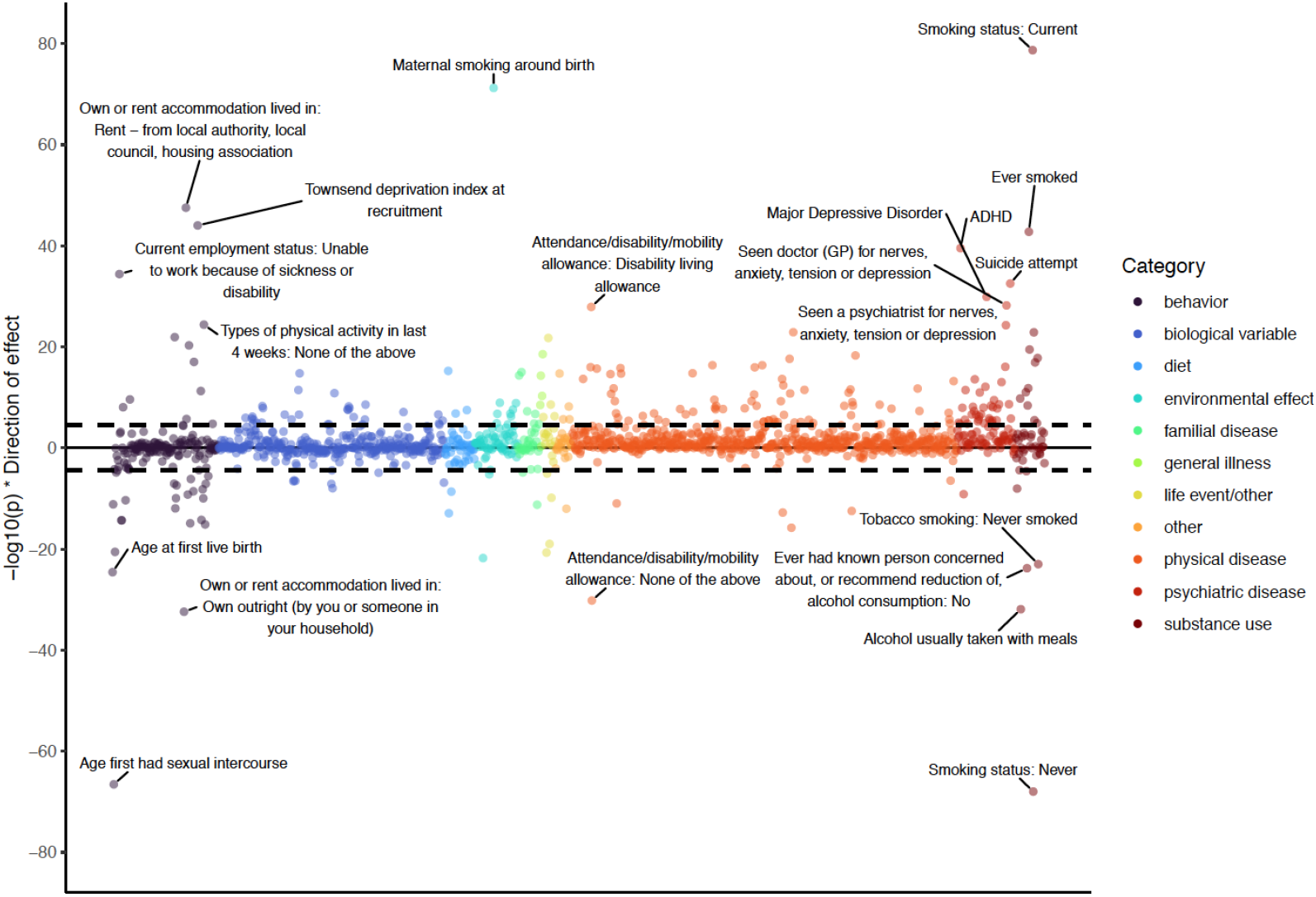
Genetic correlations between 1,547 traits and the *addiction-rf*, calculated in MASSIVE, mapped by their statistical significance (-log10(p) on the y-axis), and broad category. **The top 20 correlations are annotated**. The black dashed line represents Bonferroni significance for association (pbon= .05/1,574 = 3.232062e-05).

### Latent Causal Variable Analysis

We conducted Latent causal variable (LCV)^30^ analyses on the same 250 phenotypes that were significant in our genetic correlation analyses. In general, the genetic causality proportion (gcp, an index of the proportion of genetic causality) was nominally significant for several physical diseases (**Supplemental Table 17;** *addiction-rf* is trait 2 in all instances, therefore a negative gcp indicates a causal effect of *addiction-rf* on another trait). After correction for multiple comparisons (p = 0.05/250 = 1.98e-4), the only significant causal processes were medication codes related to cholesterol, blood pressure, diabetes and exogenous hormones where there was support for a causal role of *addiction-rf* on the physical diseases indexed by these codes.

### Polygenic risk score (PRS) analyses

#### PRS analyses with similar measures of addiction-rf and individual SUDs

In the independent Yale-Penn 3 sample^16^ (EUR n=1,986), the *addiction-rf* PRS was significantly associated with a phenotypic factor underlying several SUDs (p<.001), polysubstance use disorder (p<2e-16), and each individual SUD (DSM-IV^31^: AUD, CUD, OUD, tobacco dependence, or TD, and cocaine use disorder, or CoUD; **Figure 4; Supplemental Table 18**). Nagelkereke’s r^2^ values ranged from 2.4% for CUD to 5.9% for TD, and 6.6% for a phenotype similar to *addiction-rf* that represents phenotypic commonality across AUD, CUD, OUD, TD and CoUD. Odds ratios varied from 1.41 for CUD to 1.73 for OUD.

**Figure 4.**
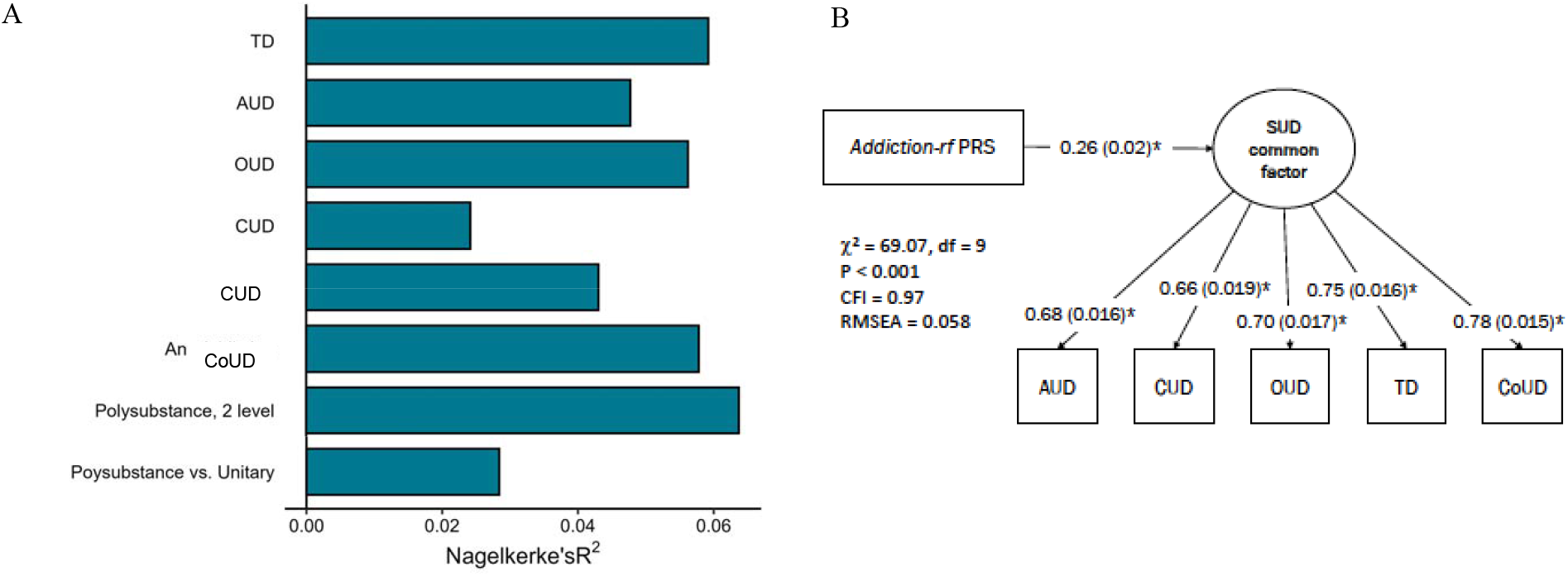
(A) Polygenic risk score (PRS) of the *addiction-rf* predicts lifetime alcohol (AUD), cannabis (CUD), opioid (OUD), tobacco (TD), and cocaine (CoUD) use disorders, and variables representing more than one lifetime substance use disorder diagnosis vs no SUDs diagnosis (Polysubstance Use Disorder, 2 level), more than one lifetime diagnosis vs. one lifetime diagnosis (polysubstance vs. unitary), as well as any substance use disorder diagnosis (Any Addiction) in an independent sample (Yale Penn 3; N=1,986 individuals of European genetic ancestry). (B) The *addiction-rf* PRS was associated with a comparable phenotypic substance use disorders (SUD) common factor in the Yale-Penn sample. Controlling for age, sex and 10 genetic principal components of ancestry, all path estimates are fully standardized. Estimates were significant at p < .001 (minimum available in LAVAAN).

#### PheWAS in Electronic Health Record data

In the BioVU sample (EUR N=66,914)^22^, the *addiction-rf PRS* was associated with SUDs (p=3.31e-29; **Figure 5**), various types of substance involvement [e.g., Tobacco Use Disorder p=9.79e-24, alcoholism (so named in EHR, we note the term “alcohol use disorder” is more appropriate), p=1.12e-21), chronic airway obstruction (p=4.99e-10)], and several psychiatric disorders, with the strongest being bipolar disorder (p=2.44e-11). Controlling for any SUD diagnosis to account for causal effects found similar associations with alcoholism, mood disorders, respiratory disease, and heart disease (**Supplemental Figure 7A**). Controlling for tobacco use disorder diagnosis did not significantly modify associations (**Supplemental Figure 7B**).

**Figure 5.**
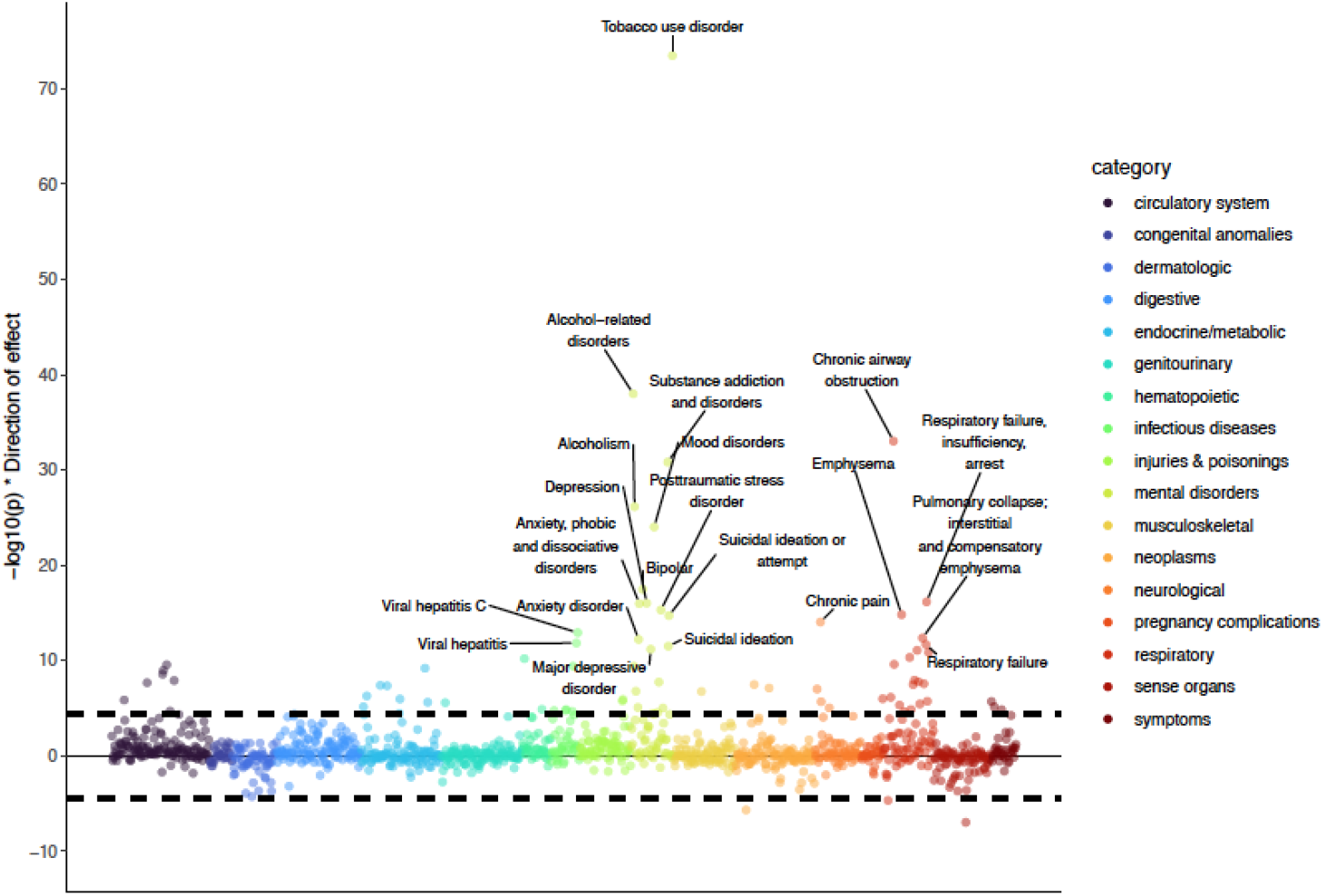
Phenome Wide Association Study (PheWAS) of the *addiction-rf* polygenic risk score (PRS) in the BioVU electronic health record database (N=66,915). Results are shown by level of statistical significance (-log10(p) of association on y-axis) and in broad categories. Analyses were conducted using logistic regression, controlling for median age on record, reported gender, and first 10 genetic ancestry PCs. The dashed lines represent Bonferroni correction for 1,335 phecodes (pbon=3.745e-05). The top associations are annotated. Symptoms represents phenotypes’ like back pain, loss of height, muscle weakness etc that are not indictive of a disease alone.

#### Behavioral and Neural Phenotypes in Substance-Naïve Children

Among 4,491 substance-naïve children aged 9-10 years who completed the baseline session of the ABCD Study^®23^, the *addiction-rf* PRS was correlated (after Bonferroni correction) with impulsivity (p=2.09e-05), family history of drug addiction (p=7.04e-07), family history of hospitalization due to mental health concerns (including suicide; p=4.64e-06), childhood externalizing behaviors (e.g., antisocial; p=1.62e-05), childhood thought problems (p=3.51e-06), sleep disturbances (p=1.52e-07), parental externalizing and substance use behaviors (e.g., prenatal smoking; p=2.87e-11), maternal pregnancy characteristics (e.g., urinary tract infection during pregnancy, p=2.70e-7), socio-economic disadvantage (e.g., child’s neighborhood deprivation; p= 9.84e-07), and child’s likeliness to play sports (p=2.80e-06) (**Figure 6**; **Supplemental Table 19** for results from all phenotypes).

**Figure 6.**
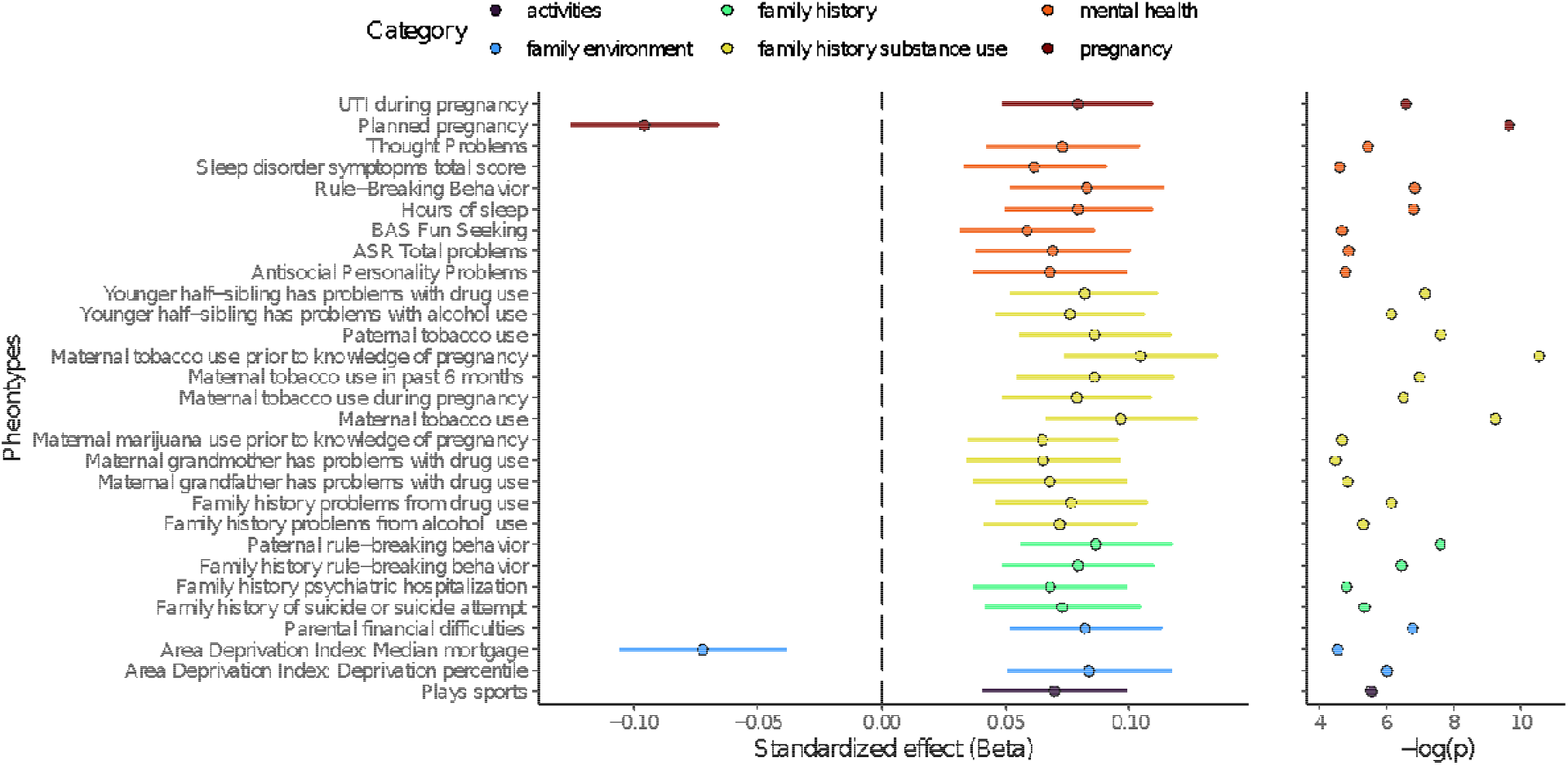
Phenome Wide Association Study (PheWAS) of the *addiction-rf* polygenic risk score (PRS) in the Adolescent brain and Cognitive Development (ABCD® 2.0) dataset (N=4,491 of European genetic ancestry). Results surviving a Bonferroni correction for 1,480 behavioral (pbon==.05/1480 = 3.378378e-05) are shown by effect size and direction with corresponding levels of statistical significance, and in broad categories. Analyses were conducted using a mixed effects model and controlled for first 10 PCs, age, sex, age by sex and random effects for family and site. ASR = Adult self-report from parents on parents symptoms.

## Discussion

Our multivariate GWAS (total n=**1,025,550**) revealed that a general liability to addiction to alcohol, tobacco, cannabis, and opioids is characterized by a highly polygenic architecture. The *addiction-rf* correlated with other medically relevant traits and illnesses. Substance-specific genetic factors with relatively less polygenic architectures, and in some instances, a single SNP of larger effect (e.g., *ADH1B* for alcohol) were also identified. Outlining this common genetic factor - *addiction-rf* - suggests that current substance-specific diagnostic classification schemes (e.g., DSM) only partially represent the phenomenon of SUD at the genetic level of analysis.

We found 17 genomic loci significantly associated with *addiction-rf*, and 27 substance-specific loci. Post-hoc fine-mapping, annotation, and exploratory drug repurposing analyses highlight the potential therapeutic relevance of the discovered loci. The *addiction-rf P*RS was associated with many medical conditions characterized by high morbidity and mortality rates, including psychiatric illnesses, self-harming behaviors, and somatic diseases that could be consequences of chronic substance use (e.g., chronic airway obstruction) or precursors to heavy substance use (e.g., chronic pain). Finally, in a sample of drug-naïve children, the *addiction-rf* PRS was correlated with impulsivity and externalizing behaviors, a family history of hospitalization for and diagnosis of severe mental illness, thought problems, and neighborhood deprivation, which reflects the shared genetic and gene-environment correlations with phenotypes and exposures present early in life.

Our analyses suggest that the regulation/modulation of dopaminergic genes, rather than variation in dopaminergic genes themselves, is central to general addiction liability. *DRD2* was the top gene signal, which was mapped via chromatin refolding, suggesting a regulatory mechanism. The role of striatal dopamine in positive drug reinforcement is well established^32^. *DRD2* plays a role in reward sensitivity and may also be central to executive functioning^33^ – the interplay of reward and cognition is likely relevant throughout the course of addiction. These complementary observations reinforce the role of dopamine signaling in addiction^34,29^. Our results give some evidence that chromatin refolding plays a role in these processes.

Other regulatory effects on dopaminergic pathways were supported by the signal at *PDE4B,* which has been implicated in prior GWASs of disinhibition traits^35^. The PDE4 antagonist, ibudilast, has been shown to reduce heavy drinking among patients with AUD^36,37^ and also shown to reduce inflammation in methamphetamine use disorder^38^. The phosphodiesterase (PDE) system has been proposed as a dopaminergic regulation mechanism, and PDE inhibitors have been tested as treatments for cognitive impairments in schizophrenia that arise from antipsychotic antagonism of dopamine receptors, albeit with side effects^39^. Further, animal studies suggest that the PDE system is associated with down-regulation of drug-seeking behaviors across opioids, alcohol, and psychostimulants^40^, convergent evidence of the potential translational significance of this gene system for therapeutics^41^.

The *addiction-rf* PRS was associated with general and specific SUD liability in an independent sample. The *addiction-rf* PRS predicted ∼6% of OUD variance, which is nearly half the total SNP-heritability of OUD^16^. The *addiction-rf* PRS also predicted variance in CoUD - as CoUD was not included in the development of the *addiction-rf* (lack of well-powered CoUD GWAS), these findings highlight the generalizability of the *addiction-rf* beyond alcohol, tobacco, cannabis and opioids. Further, the *addiction-rf* PRS explained appreciable variance for a phenotypic factor comprised of AUD, CUD, OUD, TD and CoUD (R^2^ = 6.6%), suggesting that combining information across SUDs may be a useful strategy in PRS analyses of addiction.

Considering how readily many of the top hits and mechanisms can be translated into medication targets (e.g., *PDE4B*), we conducted a reverse transcriptional drug repurposing analysis of *addiction-rf*. Rather than focusing on individual gene targets, this method uses a transcriptional profile to predict the top drugs that reverse the transcriptional pattern that we derived from a transcriptome-wide association analysis. We found >100 potential perturbagens that partially reverse the transcriptional patterns underlying general addiction risk (**Supplemental Table 17**). Some of these drugs have existing evidence for treatment of SUDs, such as varenicline, an efficacious smoking cessation drug^42^ that may also reduce craving for alcohol in smokers and drinking in non-smokers^43^. Other drugs identified in the repurposing analysis that are being actively investigated include mifepristone, which is currently under clinical investigation for the treatment of alcohol dependence^44,45;^ riluzole, for which there is early evidence of a treatment effect in stimulant dependence^46,47;^ and reboxetine, that may have utility in the treatment of CoUD^48^.

Substance-specific genetic signals fell primarily into three broad categories: drug-specific metabolism (e.g., *ADH1B* for PAU), drug receptors (e.g., *CHRNA5* for PTU, *OPRM1* for OUD), and general neurotransmitter mechanisms (e.g., *CACNA1A* for CUD). In particular, alcohol-specific effects associated with *ADH1B* and other alcohol dehydrogenase genes^49^ and tobacco-specific effects associated with *CHRNA5* and acetylcholine receptor genes were an order of magnitude greater than those noted for the *addiction-rf*. Surprisingly, even after accounting for the *addiction-rf,* dopaminergic genes (*DBH* and *PDE1C* in particular) were implicated in substance-specific effects for tobacco (PTU). On the other hand, CUD-specific genes did not include well-studied receptor targets (e.g., *CNR1*) or metabolic mechanisms (e.g., Cytochrome P450 genes). However, the cannabis-specific loci were eQTLs for *CHRNA2,* and there is some evidence that *CNR1* and *CHRNA2* expression are anti-correlated in some brain regions^50^. It is also likely, given the high loading (0.74) of CUD on *addiction-rf*, that current GWASs of CUD capture genetic vulnerability that is relatively non-specific. Alternatively, effect sizes for cannabis-specific variants may be too small to detect in the available data.

Genome-wide genetic correlations and PheWAS across multiple different samples supported the role of pleiotropy and/or causality in the relationship between problematic substance use and other disinhibitory behaviors across the lifespan. Age at first sexual intercourse was amongst the phenotypes most highly genetically correlated with *addiction-rf*. Even in predominantly substance-naïve youth, the *addiction-rf* PRS was associated with externalizing behaviors, which reinforces the developmental role of these behaviors in the etiology of drug use. Despite the genetic overlap between the *addiction-rf* and a recent index of externalizing behaviors (rG=0.63)^35^, a significant portion of the variance in *addiction-rf* was distinct.

Our analyses highlight the robust genetic association of *addiction-rf* with serious mental and somatic illness. The *addiction-rf* PRS was more strongly associated with using drugs to cope with internalizing disorder symptoms (anxiety, depression; rg=0.60-0.62) than with the individual psychiatric traits and disorders themselves (rg=0.3), suggesting that genetic correlations between SUDs and mood disorders may partially be attributable to a predisposition to use substances to alleviate negative mood states (“self-medication”)^51^. Self-harm as indexed by the ingestion of medication, suicide attempt, and family history of suicide attempt were also highly correlated with the *addiction-rf* (rg range: 0.45-0.62). The findings regarding personal and family history of suicidal thoughts and behaviors are consistent with other recent reports that addiction is more highly correlated with suicidality than most other behaviors^52-54^. As with internalizing disorders, although the observed genetic correlation could reflect a causal pathway from SUD to suicidal thoughts and behaviors^55^, latent causal variable analysis revealed a non-significant causal effect (p=0.72, **Supplemental Table 18**).

The PheWAS also provided insight into potentially complex mechanisms of genetic liability to environmental pathways of risk. In addition to indices of socio-economic status (SES), the *addiction-rf* was correlated with maternal smoking during pregnancy and ADHD, in line with evidence that effects ascribed to the prenatal environment may also be mediated by the inheritance of risk loci^56,57^. The *addiction-rf* PRS was associated with a family history of serious mental illness, which likely represents an amalgam of genetic and environmental vulnerability^58^. Finally, disability and SES were also associated with polygenic risk, further supporting the association between environmental risk factors and common genetic effects on SUD liability^9,59,60^.

This study has limitations. First, our GWAS in individuals of African-ancestry had few discoveries, underscoring the need for systematic data collection on SUDs in globally representative populations. Still, we chose to analyze and present these data as their exclusion only furthers disparities in genetic discoveries. Second, although we discovered many loci, they accounted for only a small proportion of the total variance. More samples, particularly those in diverse populations, and the integration of rarer variants are needed to discover the biological pathways that fall below genome-wide significance or are missed in GWAS. Finally, the authors note that despite interesting correlations between the *addiction-rf* PRS and a variety of traits, effect sizes were not of a clinically-relevant magnitude nor were analyses designed to make inferences that could apply in a clinical setting. Furthermore, some of the observed associations (e.g., in the ABCD® study where we examine a host of parental characteristics using offspring PRS) are likely confounded by other factors (e.g., SES), which are beyond the scope of detailed study here, and hence have no application in clinical prognostication. Critically, our analyses clearly demonstrate that genetic risk to addictions is polygenic and comprised of very small effects of many loci, and that this genetic vulnerability is accompanied by environmental risk and resilience factors. Therefore, utilizing these PRS alone to forecast outcomes in developing youth is unlikely to be of utility in the absence of rigorously conducted follow-up studies. Consequently, the authors strongly caution against using the findings from this study, including the PRS, beyond research, and particularly across ancestral groups where they can generate spurious findings if not appropriately applied.

## Conclusion

The polygenic effects of loci associated with problematic substance use, both those conferring general addiction liability and those that are substance-specific, indirectly pose risks for a range of serious medical conditions across the lifespan. However, unlike many chronic conditions (e.g., type 2 diabetes, hypertension), efficacious treatments for addictions are not widely prescribed. Large-scale GWAS could identify additional genes and biological pathways (e.g., *PDE4B* and the PDE system) that could be leveraged for medications development. Although there are independent genetic contributions to individual SUDs, this study strongly reinforces the idea that a common and highly polygenic genetic architecture underlies multiple SUDs, a finding that merits integration into medical knowledge on addictions.

## Data Availability

Data Availability: The MVP summary statistics were obtained via an approved dbGaP application (phs001672.v4.p1). The authors thank Million Veteran Program (MVP) staff, researchers, and volunteers, who have contributed to MVP, and especially participants who previously served their country in the military and now generously agreed to enroll in the study. (For details, see https://www.research.va.gov/mvp/ and Gaziano, J.M. et al. Million Veteran Program: A mega-biobank to study genetic influences on health and disease. J Clin Epidemiol 70, 214-23 (2016)). This research is based on data from the Million Veteran Program, Office of Research and Development, Veterans Health Administration, and was supported by the Veterans Administration (VA) Cooperative Studies Program (CSP) award #G002.
Publicly available data were also taken from the psychiatric genomics consortium: https://www.med.unc.edu/pgc/, and the GSCAN consortium: https://conservancy.umn.edu/handle/11299/201564

