## Supplementary material for "Multivariate genome-wide association meta-analysis of over 1 million subjects identifies loci underlying multiple substance use disorders": Methods: Methods_Submission_Final.docx

**Summary statistics from each SUD-related GWAS**

Summary statistics from the largest available discovery GWAS were used to represent genetic risk for each construct. These include **four measures of *problematic substance use or substance use disorder (SUD):*** Problematic alcohol use^1^, Problematic tobacco use^2-4^, Cannabis use disorder^5^; Opioid use disorder^6^ (described below). All GWAS summary statistics were filtered to retain variants with minor allele frequencies > 0.01 and INFO score > 0.90 for GSCAN and PGC^3,5^ and INFO score > 0.70 for the MVP^6,8^.

For the current cross-trait GWAS, we maintained the same QC metrics and only analyzed SNPs that were present in all four input GWAS, i.e., variants that passed QC thresholds at all levels, resulting in 3,513,381 SNPs in samples of European ancestry and 5,303,643 SNPs in samples of African American ancestry. The LD scores used for the genomic structural equation modeling (GenomicSEM – in European ancestry individuals only) analyses were estimated from the European sample of 1000 Genomes and only used in the European sample. We restricted analyses to HapMap3 SNPs as these tend to be well imputed and produce accurate estimates of heritability. We used the effective N, which was estimated for each GWAS^7^. For case control GWAS, the effective N was calculated as the population prevalence, while for binary GWAS we used the equation: N_effective_ = 4/((1/N_case_) + (1/N_control_))^8^ while continuous and quasi-continuous traits used the given N or if, from MTAG, using the equation N_effective_ = ((Z/$\beta$)^2^)/(2*MAF*(1-MAF))^1^.

*Problematic Alcohol Use:* Summary statistics for problematic alcohol use (PAU)^1^ were derived from a meta-analysis (in METAL)^9^ of GWAS of DSM-IV alcohol dependence from the Psychiatric Genomics Consortium^8^ (PGC-AD; n = 11,569 case, 34,999 controls), ICD-9/10 based diagnoses of alcohol use disorder from the Million Veteran Program phase 1 and 2 data (MVP; n = 45,995 cases; 221,396 controls), and the Problem subscale score from the Alcohol Use Disorders Identification Test (AUDIT-P) from the UK Biobank (n = 121,604)^10^. The case-control data were analyzed using logistic regression in MVP, in the PGC-SUD meta-analysis using logistic regression, logistic mixed models and generalized estimating equations, and in the UK Biobank (for the continuous score) using BGENIE (https://jmarchini.org/bgenie/). The final GWAS summary statistics included data on 435,563 participants. Effective N in both the MVP and PGC-SUD analyses was computed as shown above. For the quasi-continuous AUDIT symptom count, the N was the actual N per SNP. The final effective sample size (N_effective_=300,789) for LDSC was calculated by adding the effective N (N_effective_ = 179,185) from MVP+PGC to the UK Biobank N. SNP-heritability in Europeans was estimated to be 0.064. Results for African Americans were derived from a GWAS in the MVP sample (17,267 cases, 39,381 controls)^11^.

*Problematic Tobacco Use (PTU)*: We used summary statistics from the GWAS of the Fagerström Test for Nicotine Dependence (FTND) in European (N= 28,677) and African (N= 11,787) cohorts^3^. Because cigarettes per day (CPD) is an FTND item and, in European populations, is highly genetically correlated with the total FTND score (calculated rg = 0.97, CI =± 0.12)^4^, we combined CPD (from an analysis of N=263,954 individuals)^2^ and FTND into a single indicator in the European ancestry sample. We applied Multi-Trait Analysis of Genome-wide association study summary statistics (MTAG)^12^ to summary statistics for FTND and CPD to create the combined problematic tobacco use (PTU) phenotype^4^ with an effective sample size of 270,120 individuals.  Heritability was estimated at 0.072 in Europeans. For African ancestry samples, we used the FTND analysis of 9,925 individuals (CPD not available at this time for African ancestry individuals).

*Cannabis Use Disorder (CUD)*: For European ancestry data, summary statistics were derived from a GWAS meta-analysis of DSM-IV and DSM-III-R cannabis abuse and dependence from the Psychiatric Genomics Consortium (n = 5,289 cases; n = 10,004 controls), ICD-10 cannabis use disorder from the Lundbeck Foundation Initiative for Integrative Psychiatric Research (iPSYCH) (n = 2,758 cases; n = 53,326 controls), and hospital-based diagnoses from deCODE (n = 6,033 cases; n = 280,396 controls). In deCODE, adjustments for case-control imbalance and other sample-specific features was made using a genomic inflation factor. The final European-ancestry sample included 14,080 cases with CUD and 343,726 controls, with an N_effective_ = 46,351.  Heritability was estimated at 0.019 in Europeans. For African American samples, we used a GWAS meta-analysis from the Psychiatric Genomics Consortium (n = 3,848 cases, 5,897 controls)^5^.

*Opioid Use Disorder* *(OUD)*: Summary statistics were derived from a meta-analysis^6^ of GWASs of DSM-IV opioid abuse or dependence from the Yale-Penn sample and the Study of Addiction: Genetics and Environment and ICD-9/10 codes for opioid use disorder from the Million Veteran Program (individuals of European ancestry 10,544 cases; 72,163 opioid-exposed controls, N_effective_ = 30,443, African Ancestry 5,212 cases, 26,876 controls). Heritability was estimated at 0.115 in Europeans N_effective_ = 30,443.

**Genome-wide Analyses in European Ancestry individuals Using Genomic Structural Equation Modeling**

Using the summary data from individuals of European ancestry, we estimated genetic correlations between problematic alcohol use (PAU), problematic tobacco use (PTU), cannabis use disorder (CUD), and opioid use disorder (OUD). Genetic correlations ranged from 0.19 (S.E. = .04) for PAU and PTU to 0.78 (0.09) for CUD and OUD (**Supplemental** **Figures 1**). PTU showed the lowest SNP-rg with other SUD phenotypes [i.e., PAU = 0.19 (0.04), CUD = 0.31 (0.05), OUD = 0.26 (0.08)] while OUD [PAU = 0.69 (0.07) and CUD = 0.78 (0.09) showed the highest rgs]. This lower genetic correlation is discussed in our prior study^4^. The sample size of summary data derived from African American ancestry individuals was not sufficient for LD Score^13^ analyses and thus, we used a different method to analyze the African and cross-ancestry data (below).

For the European ancestry samples, we next estimated a confirmatory factor model specifying a unidimensional addiction risk factor *(addiction-rf* ) underlying the genetic covariance among PAU, PTU, CUD and OUD. GenomicSEM^14^ provides the following fit statistics: chi-square, comparative fit index (CFI), and the standardized root mean square residual (SRMR). A nonsignificant chi-square, CFI > 0.95, and SRMR < 0.08 indicate good fit for structural equation modeling^15^. CFI > 0.90 is sometimes considered acceptable fit for GenomicSEM^16^. The model fit the data well [*X*^2^(1) = .017, p = 0.895, CFI = 1, SRMR = 0.002; residual r = 0.51, p = 0.016; **Supplemental Figure 2**]. These models are described in the supplement and in our past work^4^. African American samples were not adequately powered for confirmatory factor analysis.

GenomicSEM conducts genome-wide association analyses in two stages. First, a multivariate version of LD score regression is used to estimate the genetic covariance matrix among all GWAS phenotypes, which is then combined with each individual SNP to calculate SNP-specific genetic covariance matrices. These matrices are then used to estimate the SEM using the lavaan package in R^16^. Variable and unknown extents of sample overlap across contributing GWAS are automatically accounted for in the estimation procedure. Model output from GenomicSEM is available upon request to the first author.

**ASSET: Genome-wide Analyses in African ancestry individuals, and in European Ancestry individuals to identify substance-specific SNPs**

We also ran Association analysis based on Subsets (ASSET)^17^ with data from the European and African American ancestry samples to identify variants that were only associated with one trait of interest (as well as to identify SNPs with common and specific effects in African ancestry individuals). In the European ancestry data, ASSET was used to identify SNPs with substance-specific effects, validate the SNPs with effects on the *addiction-rf* from GenomicSEM, and to meta-analyze with African ancestry results. ASSET assessed the contribution of each SNP to PAU, PTU, CUD and OUD and partialled SNPs into those with pleiotropic effects on more than one SUD (including all 4 phenotypes, although ASSET is not able to fit data to a factor model), and those with effects restricted to one SUD-related trait alone (e.g., PAU-specific SNPs). ASSET does not leverage the genetic correlation to identify variants of interest (as GenomicSEM does); instead, subset searches scaffold effects into pleiotropic and non-pleiotropic variants based on effect size and standard error derivations that estimate the degree to which the SNP-trait association is due to pooled effects across the phenotypes, vs. a single phenotype driving variant association. We designated substance-specific loci as those that were not associated with multiple traits and whose association statistics were only driven by one trait. Additionally, while ASSET does not automatically account for sample overlap, these adjustments can be made in the covariance term using genetic correlations from LDSC, which was done in our analysis.

As ASSET does not require genetic correlations (unlike Genomic SEM), we used it for the GWAS analyses in the African American datasets (as LDSC is optimized for use in European ancestry subjects alone). Therefore, results in the African ancestry data are not identical to those in the European ancestry data in that (a) any SNPs with pleiotropic effects may influence 2, 3 or all 4 SUD-related traits, (b) SNPs may have concordant or divergent direction of effect on 2 or more traits, and (c) pleiotropic effects are not assessed as influencing a common factor, such as *addiction-rf*. Therefore, the ASSET analysis of the European ancestry data also served as a comparison to the *addiction-rf* signal from Genomic SEM.

**Cross-Ancestry Meta-analysis**

We conducted a cross-ancestry meta-analysis of the European and African ancestry summary results. To be consistent across ancestries, we used ASSET results from both ancestries. In the cross-ancestry meta-analysis, we first extracted all SNPs with evidence of any pleiotropy (i.e., effects on 2 SUDs, 3 SUDs, or all 4 SUDs, including different sets of SUDs in each ancestry) from both ancestral groups to create a common set of SNPs that had some evidence of pleiotropy in both ancestral groups. A meta-analysis in METASOFT^18^ using a random-effects meta-analysis with ancestry group as a random effect was used to identify cross-ancestral effects. We calculated Cochran’s Q-statistic; any SNP with Q-statistic <5e-8, indication significant cross-ancestral heterogeneity, was removed, leaving 317,447 SNPs. We report the random effects BETA and p-value as cross-ancestry effects.

**Estimation of SUD-Specific Genetics in European Ancestry individuals: Q-SNP and ASSET One-sided P-value Test**

To validate substance-specific SNPs, we used ASSET for discovery of these variants and, in the European ancestry GWAS, also examined Q-SNP results. Q-SNP^14^ indexes violation of the null hypothesis that a SNP acts on a trait entirely through a common factor (e.g., *addiction-rf*). For example, if a SNP has a particular effect on one SUD trait (such as SNPs in *CHRNA5* influencing PTU), then it should have significant Q-SNP statistics because it violates the assumption that its effect on PTU is via the *addiction-rf*. We identified Q-SNPs by estimating the association between each SNP and the *addiction-rf*. Then, we fit a model where the SNP predicted the indicators underlying the *addiction-rf*, i.e., PAU, PTU, CUD, OUD. We compared the Chi-square difference statistic between the two models; those with significant decrement of fit (*X^2^ for Δ*df = 4) in the model where the SNP predicted the *addiction-rf* alone relative to the SNP predicting the indicators themselves was considered a significant Q-SNP above genome-wide significance (i.e. Q p < 5e-8). SNPs (which also indexed the full locus) with significant Q-SNP statistics were removed from the *addiction-rf* summary statistics for all post-hoc analyses, including fine-mapping, gene-based tests, transcriptome-wide association analyses, LD score genetic correlations and polygenic risk score analyses (e.g., PheWAS).

Q-SNP analysis also identified several SNPs that appeared to be specific to a single substance. However, as Q-SNP cannot be used for precise identification of substance-specific (trait-specific) SNPs, we relied on ASSET analyses (with a 1-sided p-value), to identify the subset of SNPs with effects (at genome-wide significance, p<5e-8) limited to only 1 SUD-related trait (e.g., PAU-specific).

**Post-hoc analyses of European ancestry GWAS results**

**Estimation of polygenicity for the *addiction-rf* vs. substance-specific genetics**

To estimate the polygenicity of the *addiction-rf* (Genomic SEM) results and the 4 input GWAS (PAU, PTU, CUD, OUD), we used a likelihood-based approach with Linkage Disequilibrium scores from the 1000 Genomes project. Effect size distributions were estimated using a flexible normal mixture model based on number of tagged SNPs and LD scores^13^.

**Biological Characterization**

FUMA^19^ was used for post-hoc bioinformatic analyses for each of the five GWAS (i.e., the *addiction-rf* (from Genomic SEM), PAU-specific, PTU-specific, CUD-specific, OUD-specific (from ASSET) loci) in only the European ancestry samples. Within FUMA, gene-based tests and gene-set enrichment were conducted via MAGMA^20^; gene annotation, and identification of SNP-to-gene associations via expression quantitative trait loci (eQTLs) or chromatin interactions (via Hi-C data) in PsychEncode^21^ and Roadmap Epigenomics tissues for Prefrontal cortex, hippocampus, ventricles, and neural progenitor cells^22,23^. In addition, we used S-MultiXcan^24^ to also conduct a transcriptome-wide association analysis across all brain tissues in the GTEx sample (v8)^25^.

**Determining Significant Lead Variants.** Because the statistical power of GWAS depends upon how well causal variants are tagged, annotation was extended beyond independent significant SNPs to incorporate all candidate SNPs. We used data from the 1000 Genomes Project (1000G)^26^ phase 3 European (EUR) population as a reference to calculate the linkage disequilibrium (LD) structure (r^2^) of pairwise SNPs and their minor allele frequencies (MAFs). Genome-wide significant SNPs (p < 5 x 10-8) that were independent (r2 < 0.1) were considered “independent significant” SNPs. All potential “candidate” SNPs refers to all SNPs available in the 1000G EUR reference panel that were in LD (r^2^ ≥ 0.6, max 1 Mb window, MAF ≥ 0.01) with the independent significant SNPs. Of the independent significant SNPs, those with the lowest p-value that were independent at r^2^ < 0.1 were defined as “lead” SNPs. LD blocks of all the identified independent significant SNPs and lead SNPs that were less than 250 kb apart were combined and characterized as genomic risk loci. Candidate SNPs were functionally annotated and used for our gene prioritization analyses, while the lead SNPs with the lowest *p*-value were used to represent their respective genomic loci.

**Fine-mapping with SusieR.** We fine-mapped the association statistics of four phenotypes (*addiction-rf*, PAU-specific, PTU-specific, CUD-specific; not enough signal for OUD-specific) that had more than 1 genome-wide significant SNP in a 1 MB region around the lead SNP to determine the 95% credible set using susieR^27^ with at most 10 causal variants (this analysis reduces the total number of SNPs at a lead genome-wide signal to those that can credibly be considered as causal SNPs). The credible set reports include the likelihood of being a causal variant; the marginal posterior inclusion probability (PIP) ranges from 0 to 1, with values closer to 1 being most likely causal.

**Functional Characterization of Lead Independent SNPs.** To determine the functional consequences of SNPs significantly associated with the *addiction-rf* and specific substances in the European ancestry sample, ANNOVAR^28^ was run in FUMA (defaults: r2 ≥ 0.6, p < 0.05, MAF ≥ 0.01) on genome-wide SNPs located within the genomic risk loci to determine their functional consequences. These SNPs were matched to ANNOVAR’s database according to location and reference and non-reference alleles and then annotated. To map candidate SNPs to genes, two different strategies were applied based on Ensembl genes (build 85) using FUMA. First, SNPs in or near (< 10 kb) protein-coding genes were positionally mapped to those genes. Second, we used FUMA’s expression quantitative trait locus (eQTL) status to annotate whether any SNPs were associated with gene expression. SNPs that significantly affect gene expression were extracted from GTEx v8^25^ and BRAINEAC^29^ samples. In FUMA, GTEx v8 gene expression data were utilized to perform eQTL mapping of the following brain tissues: amygdala, anterior cingulate cortex BA24, caudate basal ganglia, cerebellar hemisphere, cerebellum, cortex, frontal cortex BA9, hippocampus, hypothalamus, nucleus accumbens basal ganglia, putamen basal ganglia, cervical (c-1) spinal cord, and substantia nigra. The BRAINEAC database of brain-tissue-specific gene expression data was used for the following brain tissues: cerebellar cortex, frontal cortex, hippocampus, inferior olivary nucleus, occipital cortex, putamen, substantia nigra, temporal cortex, thalamus, and intralobular white matter.

To determine whether intronic independent SNPs served a possible regulatory function, significant independent SNPs were annotated within FUMA for chromatin-chromatin interaction via 3D chromatin interaction (Hi-C)^30^ in the European ancestry sample. Hi-C examines whether SNPs reflect long-range enhancer-promotor associations. Hi-C data from the following pre-existing builds were used in FUMA: dorsolateral PFC, hippocampus, and neural progenitor cells.

**Gene-based Analysis.** To determine which genes were significantly associated with *addiction-rf* and specific substances and create a prioritized list of genes based on the degree of association, MAGMA v1.6^20^ gene analysis was performed in FUMA. MAGMA combines the *p*-values of SNPs, mapped by physical position to protein-coding genes, to generate a gene-based *p*-value.

**Gene-set Analysis.** To detect biological pathways significantly associated with *addiction-rf* and specific substances, we used MAGMA to run a competitive gene-set analysis and cell-type specific gene-set analysis. This analysis accounts for potential confounding variables, such as gene density and size^31^. A competitive gene-set analysis is a gene-level linear regression model designed to determine whether genes within a gene-set have a significantly greater association (via combined p-value) with the phenotype than all other genes outside of the gene-set. Gene-sets are determined by shared biological and functional characteristics between genes defined by the datasets in MSigDB 6.1.^32^ For the cell-type specific analysis, FUMA was used to annotate our findings with quantitative trait locus (QTL) information from RNA cell-type specific studies of human postmortem cortex^33^, hippocampus^34^, and frontal cortex^35^ taken during prenatal development.

**Gene-property Analysis to Determine Tissue Specificity.** To evaluate in what tissues SNP effects across the whole genome are likely expressed, MAGMA gene-property analysis was was performed in FUMA on 30 general and 53 specific GTEx v8^30^ tissue types. Gene-property analysis calculates enrichment based on continuous overlap rather than comparisons across sets.

**Drug Repurposing**

Our signature matching technique used data from the Library of Integrated Network-based Cellular Signatures (LINCs) L1000 database^36^. The LINCs L1000 database catalogues *in vitro* gene expression profiles (signatures) from thousands of compounds in over 80 human cell lines (level 5 data from phase I: GSE92742 and phase II: GSE70138)^37^. We selected compounds that were currently FDA approved or in clinical trials (via <https://clue.io/repurposing#download-data>; updated 3/24/20). Our analyses included signatures of 829 chemical compounds (590 FDA approved, 239 in clinical trials) in five neuronal cell-lines (NEU, NPC, MNEU.E, NPC.CAS9 and NPC.TAK), a total of 3,897 signatures.

We matched *in vitro* medication signatures with *addiction-rf* signatures from the transcriptome-wide association analyses (conducted using S-MultiXcan)^24,38^ via multi-level meta-regression. Similar to the popular drug-discovery tool (<http://www.ilincs.org/ilincs/signatures/search/>), we computed weighted Pearson correlations between transcriptome-wide brain associations and *in vitro* L1000 compound signatures, weighting each gene by its proportion of heritability explained (*h*^2^_MULTI-XCAN_), using the metafor package in R.^39^ We treated each L1000 compound as a fixed effect incorporating the effect size (*r_weighted_*) and sampling variability (*se^2^_r_weighted_*) from all signatures of a compound (e.g., across all time points, cell lines, doses). Our analyses brain region as a random effect to account for any tissue specific heterogeneity. Both the genes for the transcriptome wide association analysis input and the medications from our drug repurposing analyses were required to survive a Bonferroni correction for multiple testing (transcriptome-wide correction = .05/14,389 = 3.48e-06; Perturbagen correction = .05/3,897 = 1.28e-05).

**Polygenic Risk Score Analysis in Yale-Penn**

**Yale-Penn.** The Yale-Penn^6,40^ sample includes 11,332 genotyped and phenotyped individuals recruited across three phases (i.e., Yale-Penn 1, Yale-Penn 2, and Yale-Penn 3) based on the time of recruitment and genotyping array used. All cohorts were ascertained via recruitment at substance use treatment centers or targeted advertisements for genetic studies of cocaine, opioid, and alcohol dependence, resulting in a sample highly enriched for problematic substance use, as well as control subjects and relatives. All participants were assessed using the Semi-Structured Assessment for Drug Dependence and Alcoholism (SSADDA)^41^. Analyses based on Yale-Penn 1 and 2 have been published previously^40^, including data used in the discovery sample of the present study; the use of data from Yale-Penn 3^6^ was limited to replication analyses or as a target sample for polygenic risk score analyses^43^ and are independent from the discovery GWASs. Yale-Penn 3 comprises 3,026 genotyped and phenotyped Americans of European (EUR; N=1,986) and African (AFR; N=1,040) ancestry passing standard quality control. Genotyping was performed at the Gelernter lab at Yale University using the Illumina Multi-ethnic Global Array containing 1,779,819 markers, followed by genotype imputation using Minimac3^42^ and the Haplotype Reference Consortium reference panel^43^ as implemented on the Michigan imputation server (https://imputationserver.sph.umich.edu).

For the present analysis, only Yale-Penn 3 EUR subjects (N=1,986) were included. DSM-IV^44^ substance abuse and dependence diagnoses (combined as abuse or dependence to represent use disorder) based on SSADDA assessments were used to determine case and control status for alcohol use disorder (AUD), cannabis use disorder (CUD), cocaine use disorder (CoUD), tobacco dependence (TD), and opioid use disorder (OUD). Of the 1,986 EUR subjects, 42.5% met criteria for AUD (N=843), 25.9% met criteria for CUD (N=515), 33.9% met criteria for CoUD (N=503), 31% met criteria for TD (N=615), and 22.6% met criteria for OUD (N=448). The mean age of Yale-Penn 3 EUR subjects is 41.5 (SE=15.1) and 51.5% are female (N=1,023).

We calculated the *addiction-rf* PRS using the PRS-cs auto approach^45^. This method assumes a general distribution of effect sizes across the genome, and then reweights SNPs based on this assumption, their effect size in the original GWAS, and their linkage disequilibrium (LD); weights for every SNP were then summed to create a final score. PRS were associated with phenotypes in Yale-Penn 3 via a logistic regression (method for SUD Common Factor below) controlling for first 10 PCs, age, sex and age by sex. These logistic regression analyses also examined whether there were associations with a variable representing any SUD (*Any Addiction*) vs no SUD, having 2 or more SUDs with controls defined as those with <2 SUD diagnoses (*Polysubstance.2level*), and cases defined as those with 2 or more SUDs with controls defined as those with 1 SUD (*Polysubstance.Unitary*). The association between the *addiction-rf* PRS and the SUD Common Factor was estimated with lavaan^16^ where a common factor loaded on the 5 disorders.

**Genetic Correlations and Latent Causal Variable (LCV) modeling**

To examine phenotypes that were genetically correlated with the *addiction-rf*, we calculated genetic correlations using LD score regression^13,46^ through the MASSIVE pipeline^47^, which conducts LD score regression^13,46^ and Latent Causal Variable Analysis^48^ on 1,547 summary statistics for various phenotypic traits, including a mixture of ICD codes and self-reported traits from the UK Biobank^52^ and publicly available meta-analyses from GWAS consortiums.

**Phenome-wide Association Studies (PheWAS)**

**PheWAS in adult samples.** As MASSIVE includes a fairly sparse set of diagnoses (not all ICD codes are available) for genetic correlation analyses, we conducted additional and theoretically relevant PheWASs using the *addiction-rf* PRS. We used electronic health records (EHR) data for 66,914 genotyped individuals of European-ancestry from the Vanderbilt University Medical Center biobank (BioVU)^49^. BioVU is a repository of leftover blood samples (~240,000 samples) from clinical testing, that are sequenced, de-identified, and linked to clinical and demographic data^55^. Genotyping and quality control of this sample have been described elsewhere^5,49^. The *addiction-rf* PRS was used to predict 1,335 diseases in a logistic regression model, controlling for median age on record, reported gender, and first 10 genetic ancestry PCs. For an individual to be considered a case, they were required to have two separate ICD codes for the index phenotype, and each phenotype needed at least 100 cases to be included in the analysis. We used a Bonferroni-corrected phenome-wide significance threshold of 0.05/1335=3.7E-05; this is overly conservative because it incorrectly assumes independence between phenotypes^50^.

**ABCD PheWAS of phenotypes collected in childhood.** To identify phenotypes that were associated with the *addiction-rf* before the onset of regular substance use, we used data from the Adolescent Brain and Cognitive Development (ABCD®) Study release 2.0 for genome data and 3.0 for phenotypes to conduct a phenome-wide association analysis of behavioral, social, and imaging phenotypes in adolescence. The ABCD Study is an ongoing multi-site longitudinal study of child health and development^51^. Children (N=11,875; including twins and siblings) ages 8.9-11 were recruited from 22 sites across the United States to complete the ABCD Study baseline assessment. We restricted our sample to participants of genomically-confirmed European ancestry (based on principal components) who were not missing on any covariates (N=4,490)^51,52^.

All non-imaging phenotypes from the ABCD Study’s baseline (version 2), 1-year follow up, and 2-year follow-up assessments were downloaded from NDA (https://nda.nih.gov/abcd). Specifically, we focused on variables assessing substance use and expectancies, psychopathology, family history of psychiatric conditions, prenatal substance exposure and perinatal events, family and neighborhood environment, physical health and illness, cognitive ability, and child activities (i.e., sports involvement and screen time; see all phenotypes in **Supplemental Table 21**, and **Supplemental Table 22** for a list of measures considered and included). Procedures for initially identifying variables included first sub-setting the data to only include the genomically-confirmed European ancestry sub-sample. Next, all missing values (e.g., 999, 777) were removed. Variables were then examined for frequency. Variables with fewer than 100 data points (and categorical variables with fewer than 100 participants endorsing minority categories) were excluded. Remaining variables were reviewed for: 1) relevance of variable for inclusion by authors, 2) sufficient variability for inclusion, and 3) response options for the variable. As a final check, a second researcher reviewed included variables for all of the aforementioned criteria. A total of 1,480 non-imaging phenotypes.

***ABCD Genotyping, Quality Control, and Imputation.*** We used the 2.0 release of the ABCD genetic data. The Rutgers University Cell and DNA repository genotyped saliva samples on the Affymetrix (Thermo Fisher Scientific; Santa Clara, CA, USA) Smokescreen array^53^. Genotyped calls were aligned to GRCh37 (hg19), and all individuals self-reporting ancestral origins (i.e., self-reported race) other than European were excluded because evidence that the predictive utility of polygenic risk scores suffers when applied across ancestral origins^54^. We separated individuals into self-reported populations before genotype QC. Genomic data was also used to confirm that individuals belonged to self-reported racial/ethnic groups.

The following preprocessing steps were conducted with the Ricopili pipeline^55^: SNPs with call rates ≥ 0.95 and MAF ≥ 1% were retained. Individuals with high rates of missingness (>5%) and autosomal heterozygosity deviation (FHET) outside of ± 2 SD were removed. After sample QC, SNPs were further filtered to call rate ≥ 0.98 and Hardy-Weinberg p-values > 1E-6 (founders only), which yielded 372,342 SNPs.

Individuals whose data passed the first phase of QC were then checked for relatedness (both known and cryptic) and Mendelian errors were resolved. Next, using data from unrelated individuals (pi-hat ≤ 0.20) and an LD pruned set of common (MAF>0.05) and non-palindromic SNPs (and excluding MHC and chromosome 8 inversion region), principal components analysis (PCA) was performed in EIGENSTRAT using the European 1000 Genomes Project phase 3 data. Only those individuals whose data aligned with non-Hispanic European ancestry were retained. Due to the sensitivity of the PRS approach to admixture, we took a conservative approach and performed stringent exclusion for ancestral outliers, consistent with the Psychiatric Genomics Consortium’s Ricopili pipeline. After selection, a final ancestrally-informative PCA was conducted, and the first 20 PCs were projected from founders to other relatives. Imputation to 1000 Genomes and Haplotype Reference Consortium (HRC) data for Europeans was conducted using strictly QCed SNPs on the Michigan Imputation Server. Dosage data were converted to hard-call genotypes using Plink, and only SNPs with imputation r^2^ scores ≥ 0.3 were used to create polygenic scores. PRS were generated using the PRS-cs software package^45^ in line with our polygenic prediction models in adults. Substance-specific PRS were not evaluated because these GWAS were predominantly driven by genes that influence response to each substance and this population is (beyond a few sips of alcohol) substance naïve.

Associations between *addiction-rf* PRS and phenotypes were estimated using mixed-effects models in the lme4^56^ package in R. Family ID and MRI serial number were included as random effects to account for non-independence of measurement associated with relatedness and scanner/site. We controlled for the first 10 ancestry principal components, age, sex, age by sex. We used a Bonferroni-corrected phenome-wide significance threshold of 0.05/1480= 3.38e-05; all results are presented in the **Supplement Table 20.**
