## Supplemental Figures for "Multivariate genome-wide association meta-analysis of over 1 million subjects identifies loci underlying multiple substance use disorders": Hatoum-cross-SUD_suppfigs (2).pdf

**Supplemental Figure 1.** Circos Plots from Hi-C

**Supplemental Figure 2.** Manhattan plots of substance-specific findings

**Supplemental Figure 3.** African Ancestry (AA) Multivariate GWAS Manhattan Plots

**Supplemental Figure 4.** Manhattan Plot of Cross-Ancestry GWAS for pleiotropic variants. Enrichment of Chemical and Perturbagen Gene Sets

**Supplemental Figure 5.** Polygenic Architecture of Substance Use Disorder Phenotypes.

**Supplemental Figure 6.** Enrichment of Chemical and Perturbagen Gene sets in FUMA for the transcriptome-wide association analysis (using MultiXcan) of *addiction-rf*.

**Supplemental Figure 7.** PheWAS of Addiction-*rf* accounting for (A) SUD diagnosis and (B) Tobacco Dependence in the BioVU.

**Supplemental Figure 8.** Factor Structure of 4 SUD GWAS.

Chromosome 1

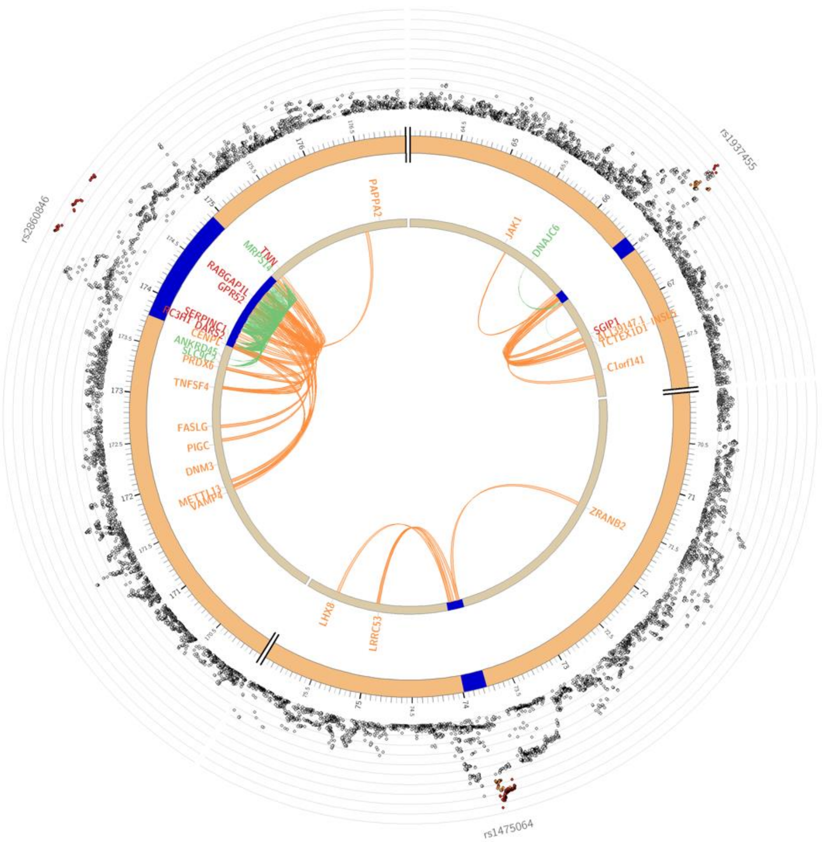

Chromosome 2

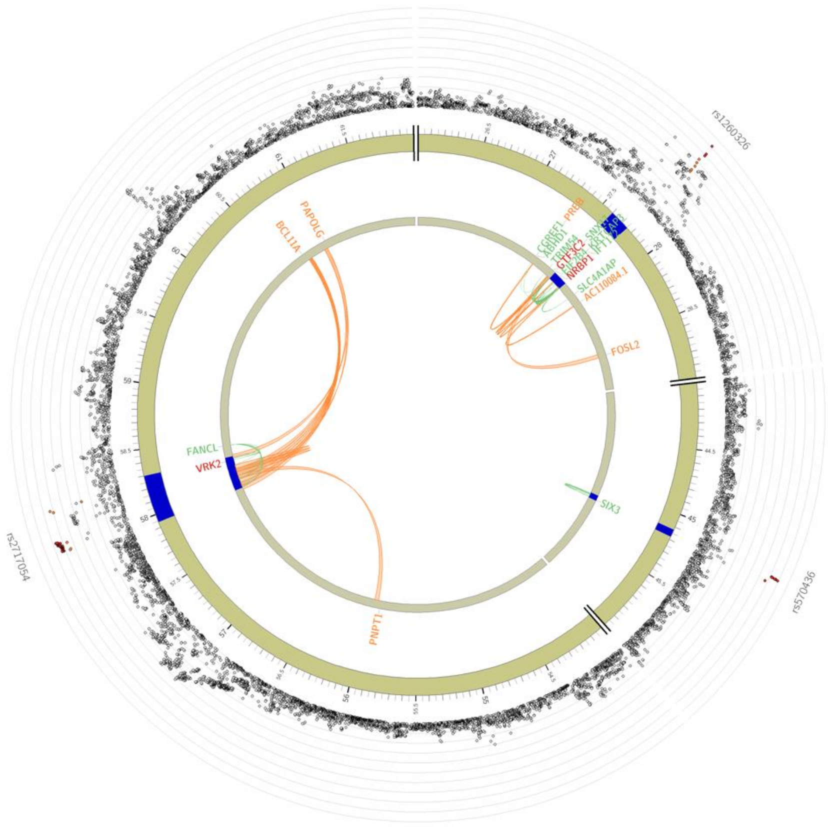

Chromosome 3

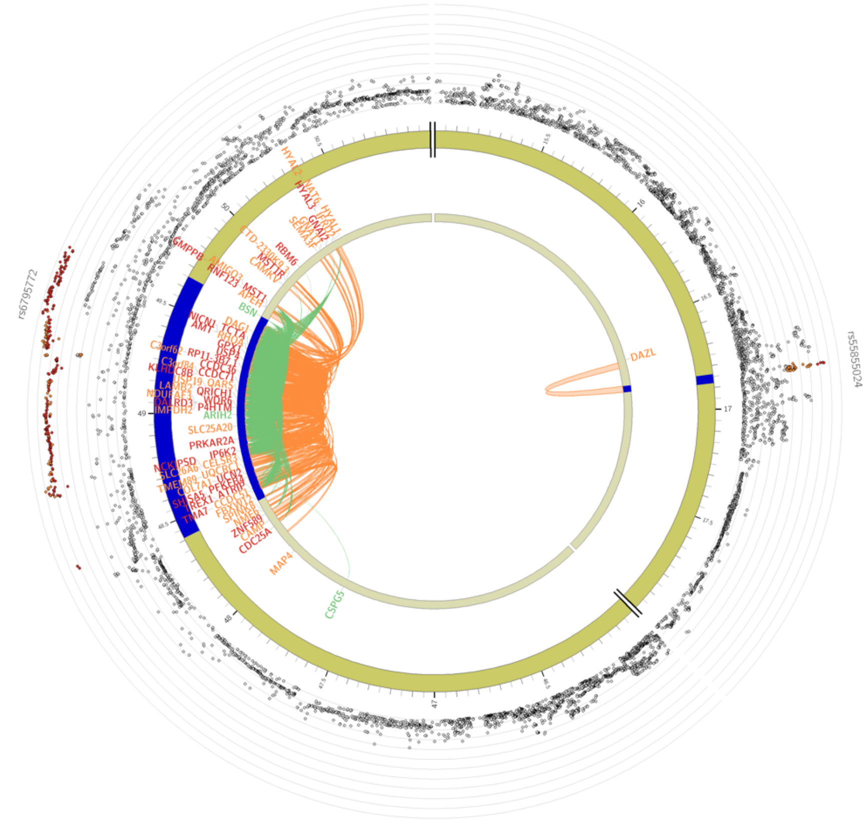

Chromosome 4

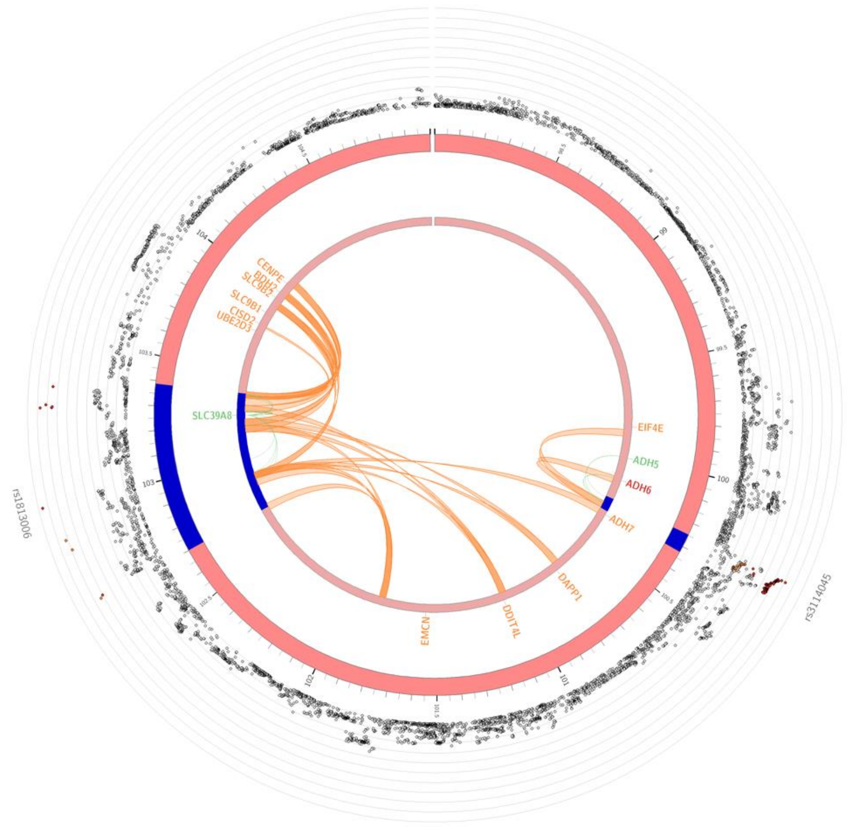

Chromosome 9

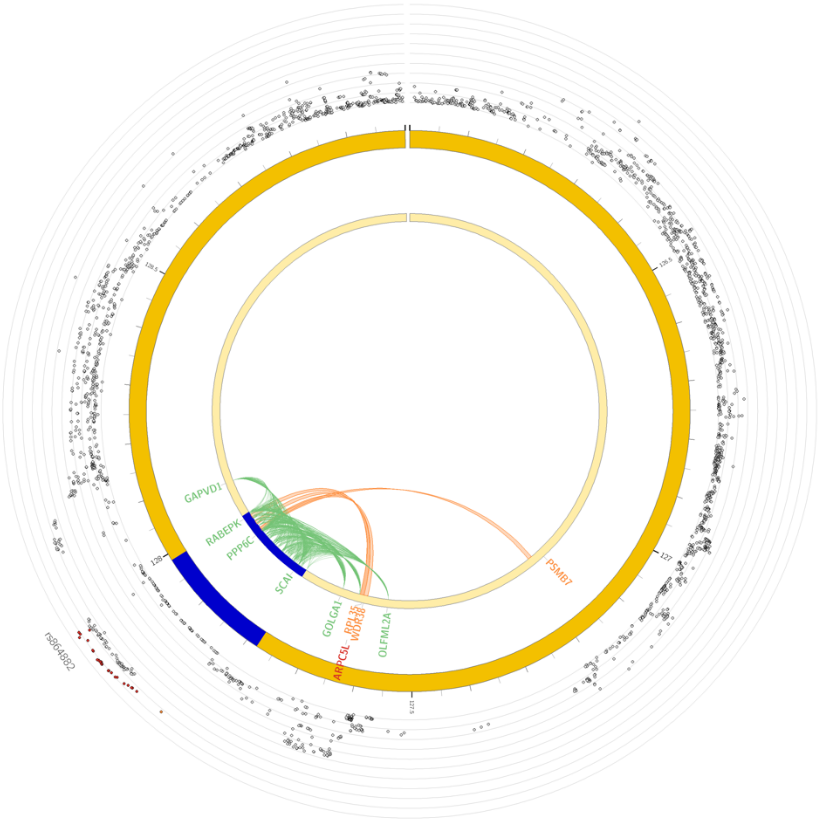

Chromosome 10

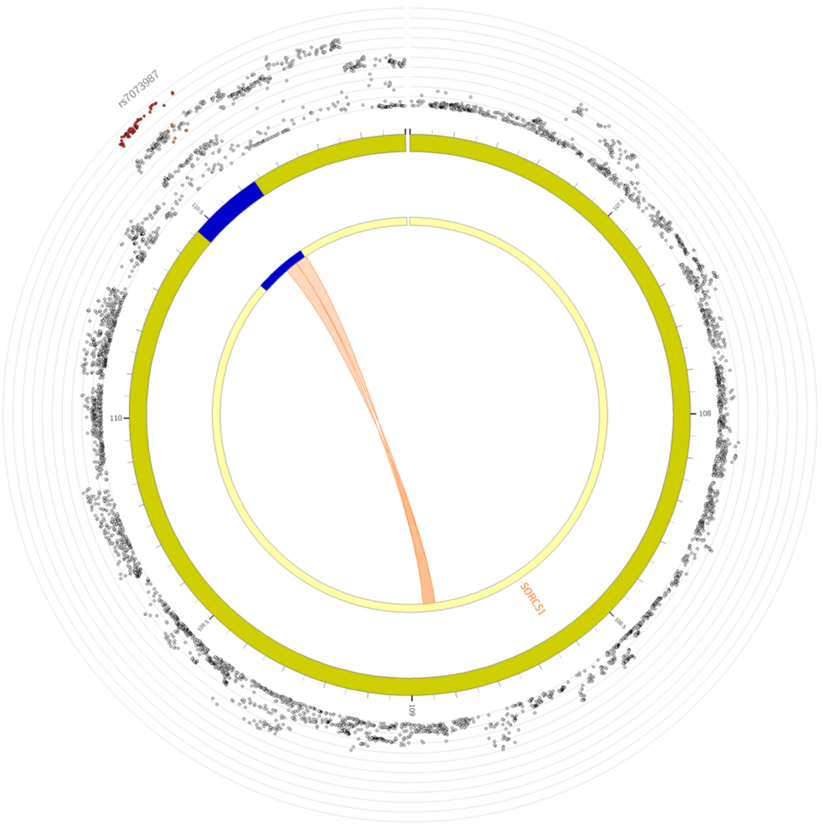

Chromosome 11

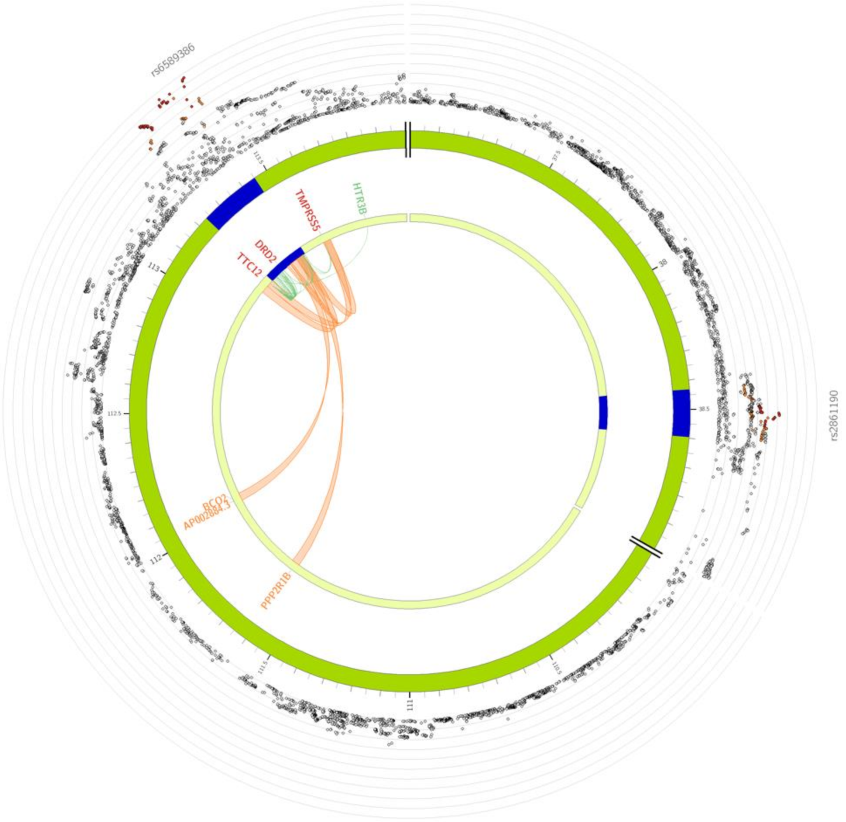

Chromosome 14

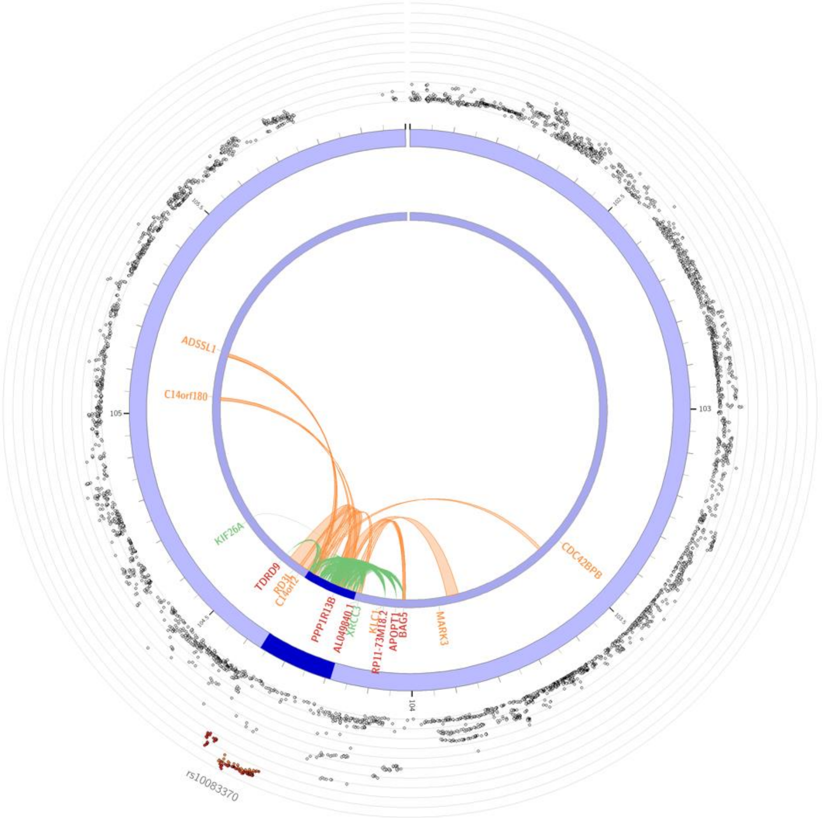

Chromosome 16

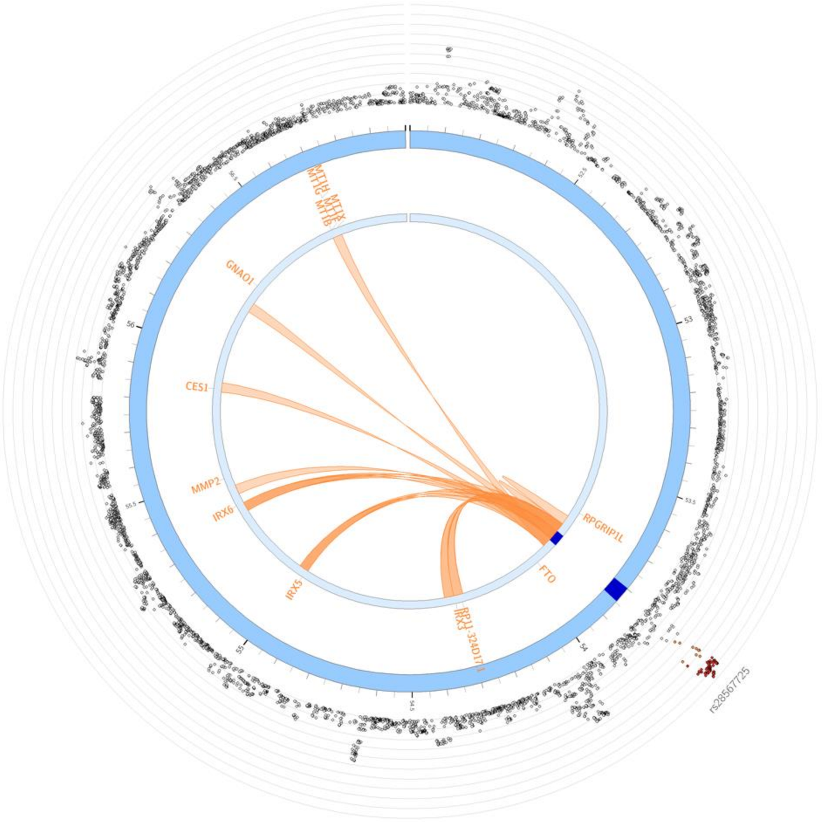

Chromosome 20

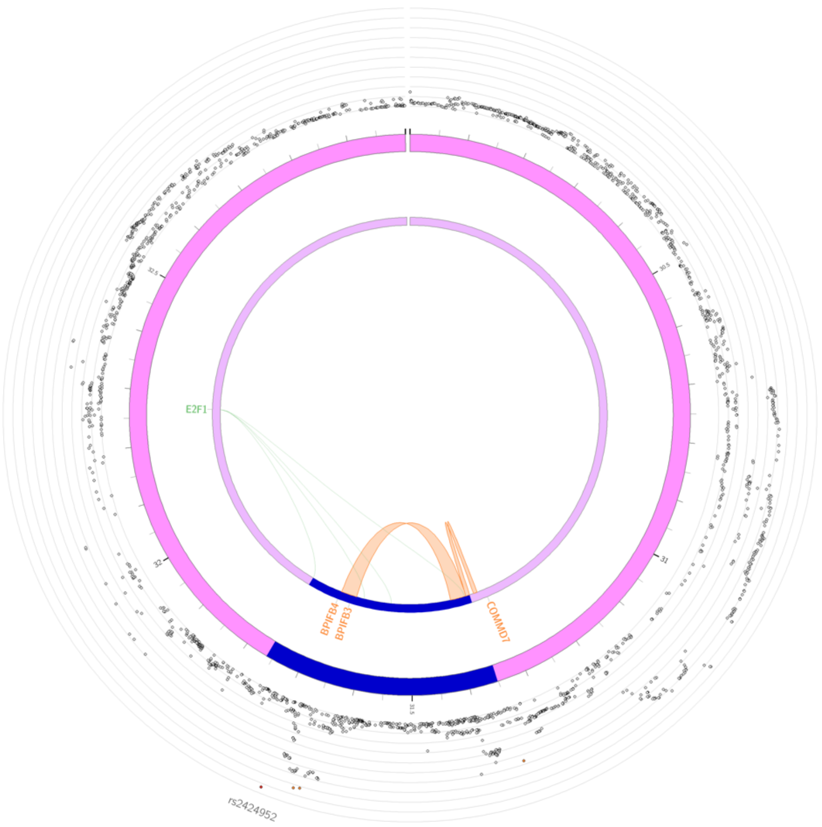

**Supplemental Figure 1. Circos Plots for chromosomes where top SNPs demonstrated chromatin contact.** Chromosome is a circle with SNPs plotted by their negative log 10 p-value (lead SNPs annotated). Orange lines represent chromatin contact with a gene. Green lines represent cis-eQTLs with a gene.

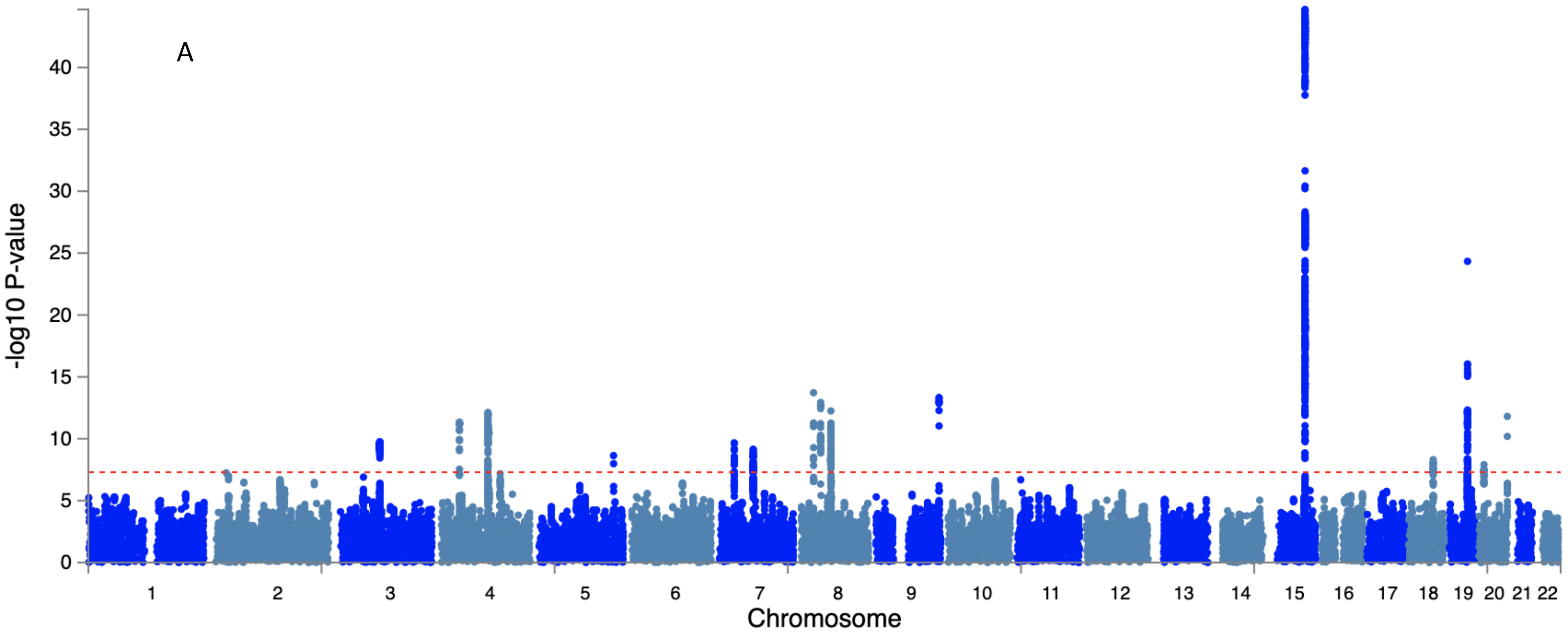

A
-log10 P-value
0
5
10
15
20
25
30
35
40
Chromosome

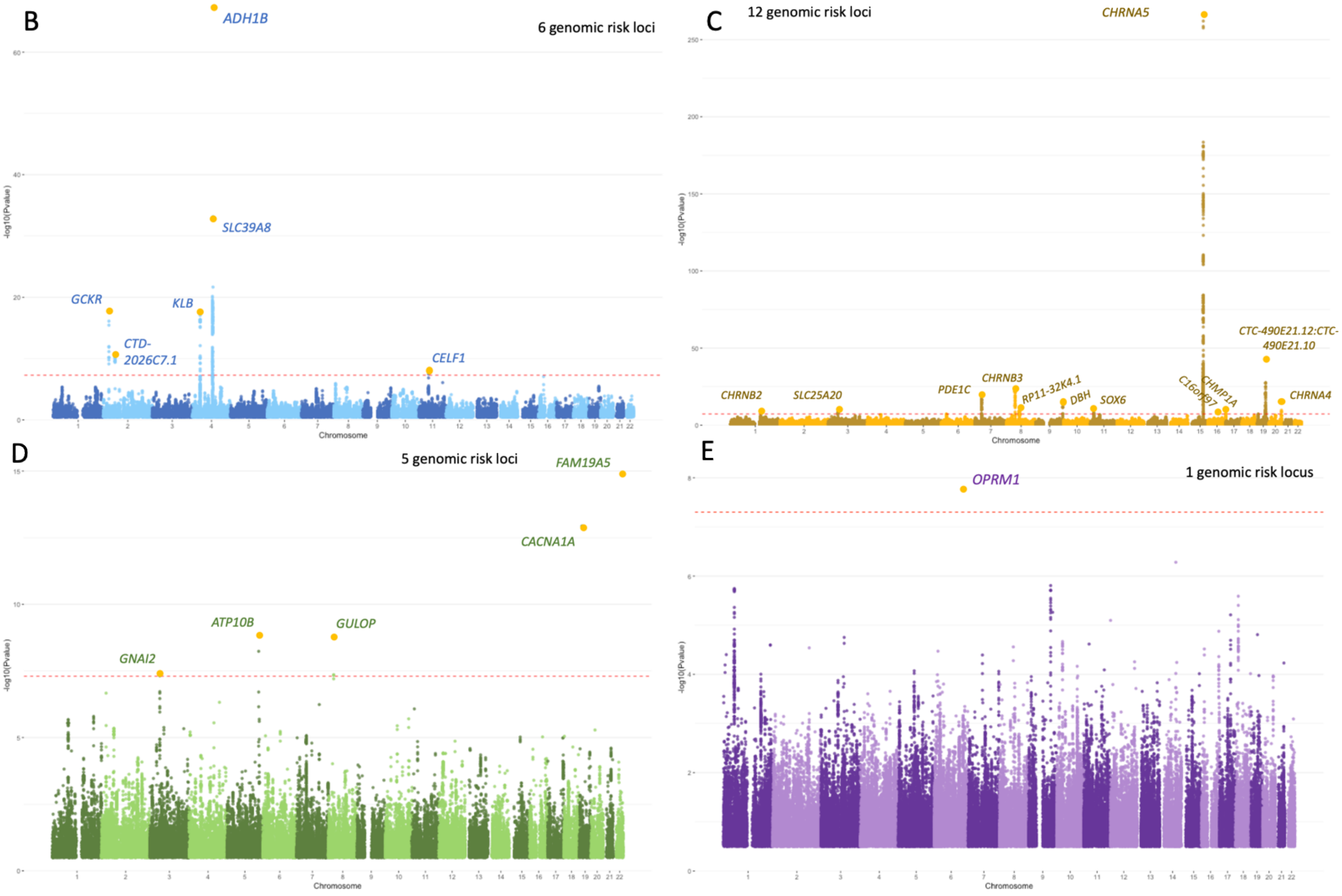
B
6 genomic risk loci
ADH1B
SLC39A8
GCKR
KLB
CTD-2026C7.1
CELF1
C
12 genomic risk loci
CHRNA5
CTC-490E21.12:CTC-490E21.10
CHRNB2
SLC25A20
PDE1C
CHRNB3
RP11-32K4.1
DBH
SOX6
CHMP1A
CHRNA4
D
5 genomic risk loci
FAM19A5
CACNA1A
ATP10B
GULOP
GNAI2
E
1 genomic risk locus
OPRM1
-log10(Pvalue)
Chromosome

**Supplemental Figure 2.** Manhattan plots of substance-specific findings. (A) Initial Q-SNP analysis p-value. Panels B-E are based on ASSET analyses. (B) SNPs associated with PAU only (C) SNPs associated with PTU only (D) SNPs associated with CUD only and (E) SNPs associated with OUD only

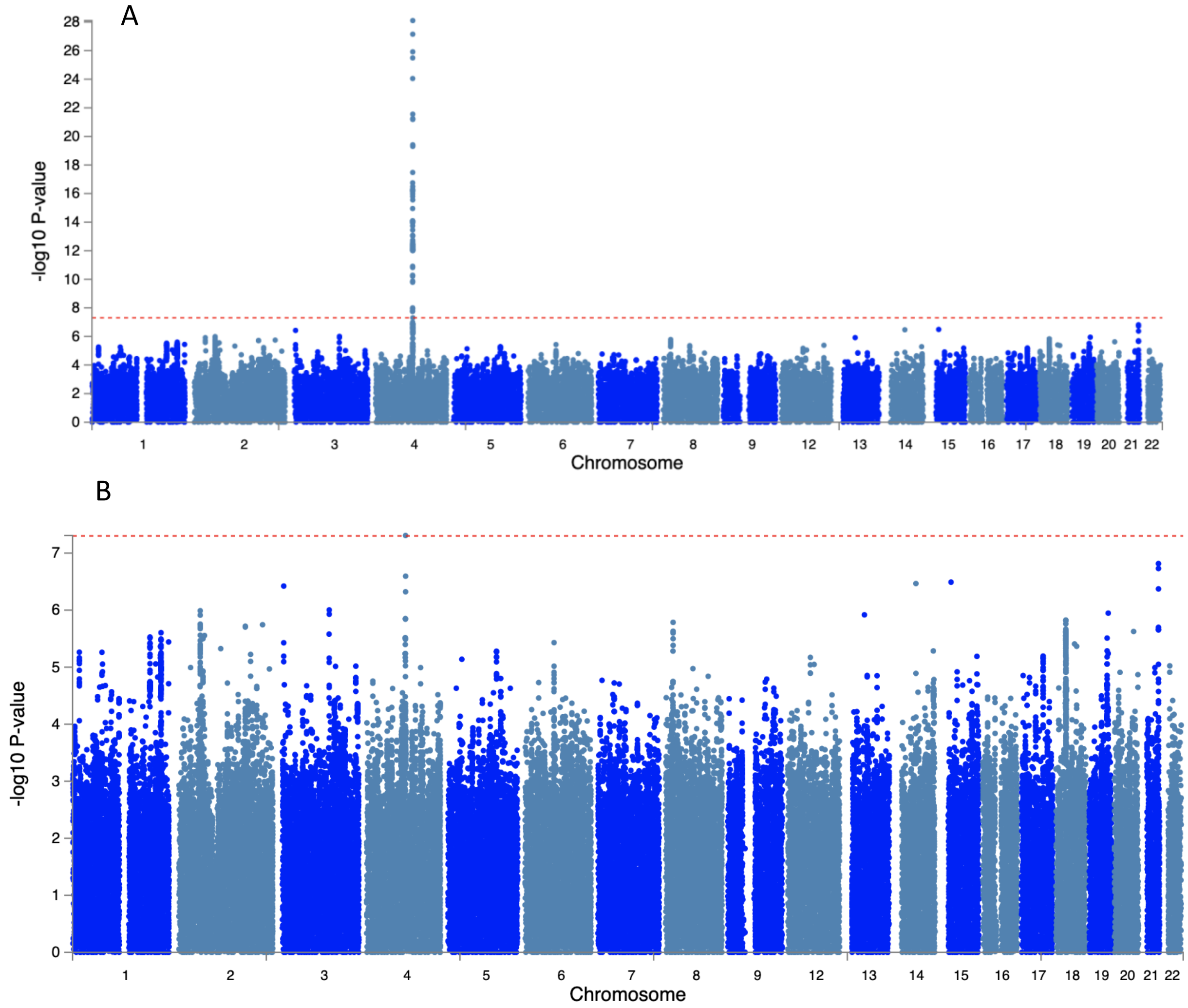
A
-log10 P-value
28
26
24
22
20
18
16
14
12
10
8
6
4
2
0
1
2
3
4
5
6
7
8
9
Chromosome
B
-log10 P-value
7
6
5
4
3
2
1
0
1
2
3
4
5
6
7
8
9
Chromosome

**Supplemental Figure 3.** African Ancestry (AA) Multivariate GWAS Manhattan Plots. **(A)** GWAS results from ASSET including all SNPs, note the significant peak on chromosome 4 that maps to ADH1B and was alcohol specific. **(B)** Loci showing evidence of pleiotropy between at least two different use disorder variables.

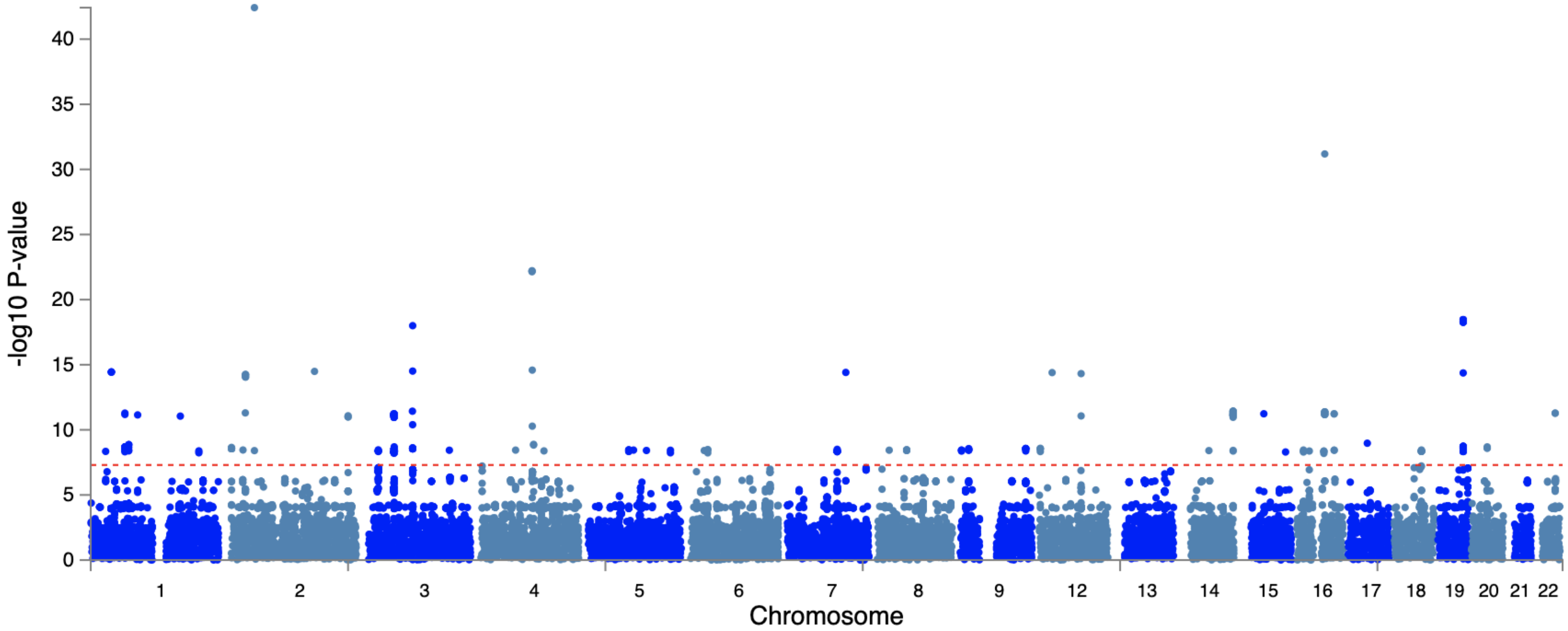

**Supplemental Figure 4.** Manhattan Plot of Cross-Ancestry GWAS for pleiotropic variants. The plot is sparse and only includes 317,447 loci that existed in both ancestral groups and showed some evidence of pleiotropy within each ancestral group.

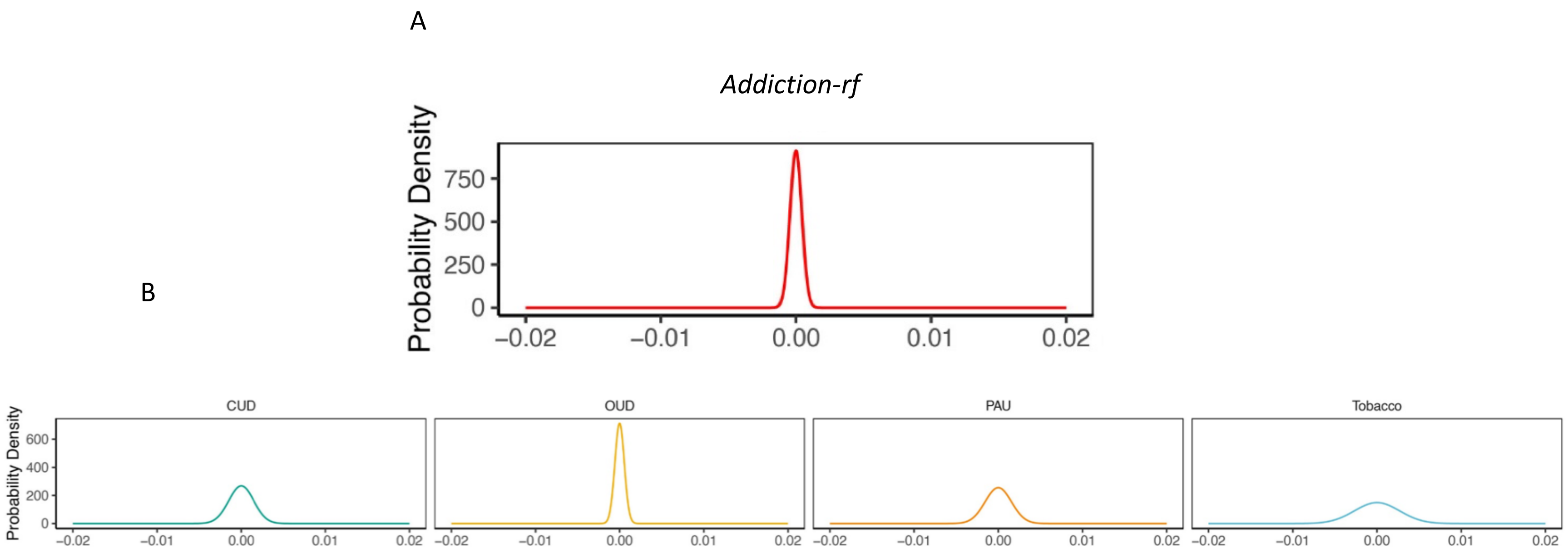

**Supplemental Figure 5. Polygenic Architecture of Substance Use Disorder Phenotypes.** Distribution of SNP effect sizes for the *addiction-rf* and substance-specific components. Taller distributions mean generally smaller effect sizes. (A) Effect size distribution for *addiction-rf*. (B) Distributions for individual substance use disorder/problematic substance use GWAS that comprised *addiction-rf*. Broadly, the *addiction-rf* had higher polygenicity. PAU = problematic alcohol use, PTU = problematic tobacco use, CUD = cannabis use disorder, OUD = opioid use disorder

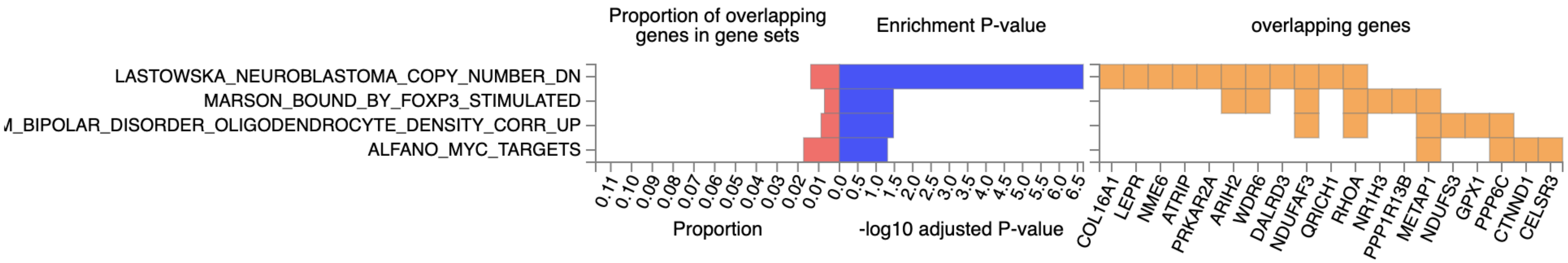

**Supplemental Figure 6.** Enrichment of Chemical and Perturbagen Gene sets in FUMA for the transcriptome-wide association analysis (using MultiXcan) of *addiction-rf*. We plotted the proportion of the genes that overlap with the total set (red), genes individually overlapping (gold), and the Bonferroni corrected -1og10 enrichment p-value for each gene set.

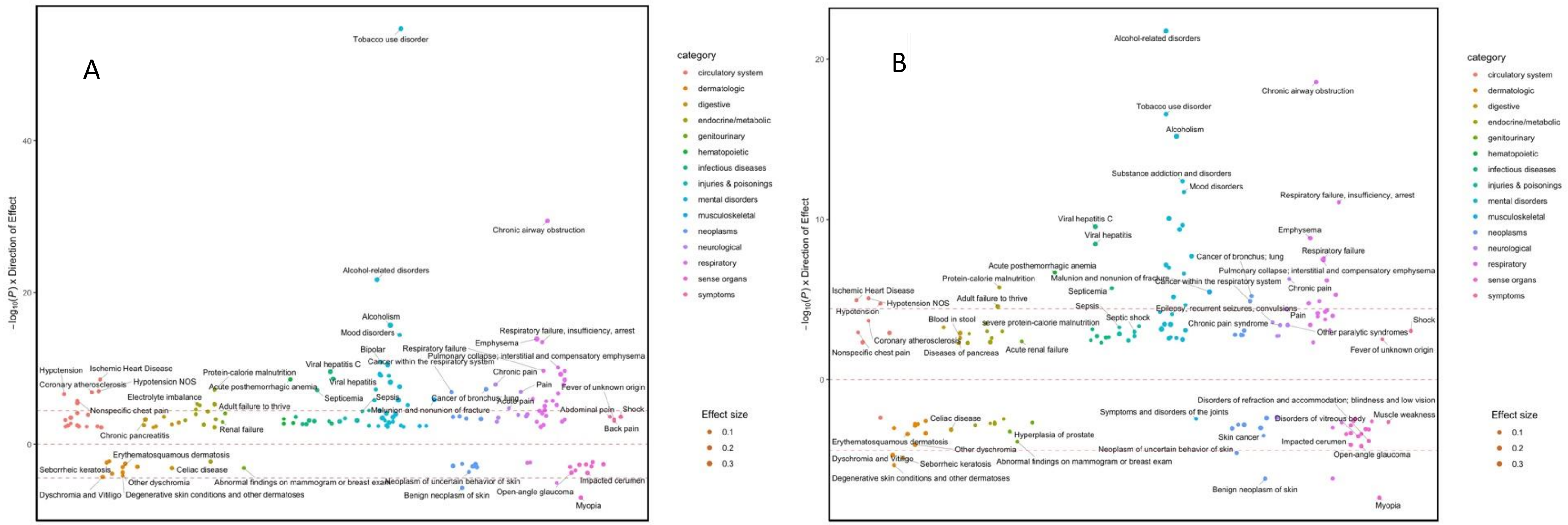

**Supplemental Figure 7.** PheWAS of Addiction-*rf* accounting for (A) SUD diagnosis and (B) Tobacco Dependence in the BioVU electronic health record database (N=66,915). Results are shown by level of statistical significance (-log10(p) of association on y-axis) and in broad categories. Analyses were conducted using logistic regression, controlling for median age on record, reported gender, and first 10 genetic ancestry PCs. The dashed lines represent Bonferroni correction for 1,335 phecodes (pbon=3.745e-05). The top associations are annotated. Symptoms represents phenotypes like back pain, loss of height, muscle weakness etc that are not indictive of a disease alone.

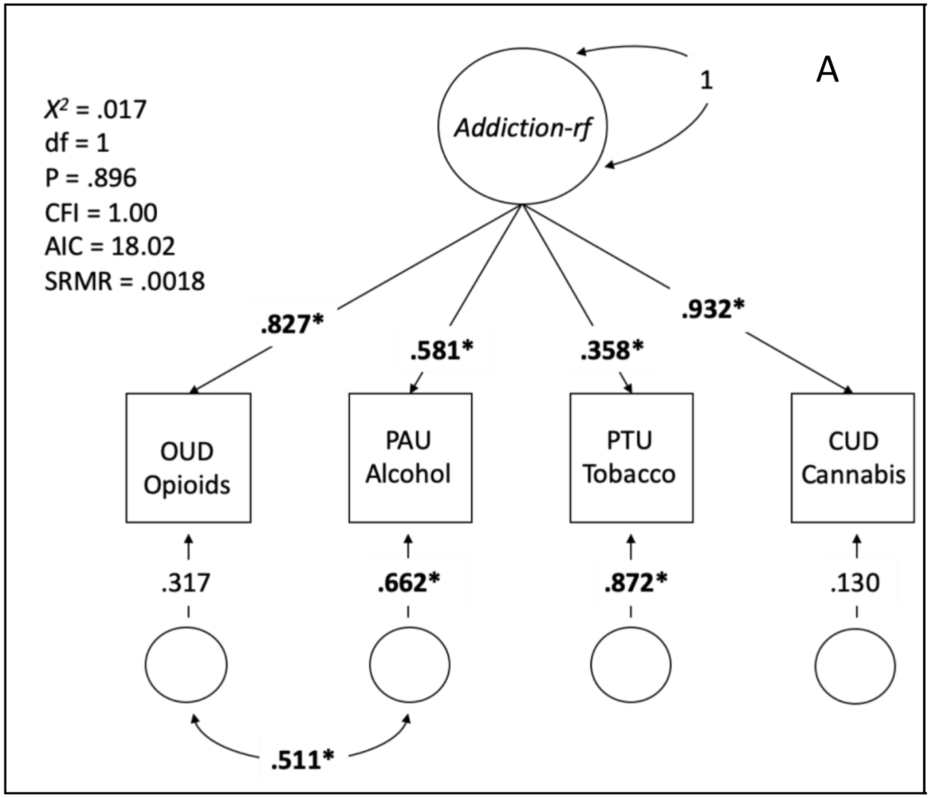

**Supplemental Figure 8. Factor Structure of 4 SUD GWAS. Factor** loadings and model fit were recreated from Hatoum et al. 2021.

We allowed all 4 SUD categories to load on a latent factor with a residual correlation between PAU and OUD was added to account for their assessment using electronic health records in the MVP cohort. *Addiction-rf=The Addiction Risk-Factor*. **Bold*** represents significance at $p < .05$.
